# Role of Dynamin 2 in mitochondrial fission and cell cycle regulation: *Dysregulation of a miR-124-3p-STAT3-DNM2-Drp1-RGCC pathway connects fission and cell proliferation in pulmonary arterial hypertension*

**DOI:** 10.1101/2024.12.05.24318153

**Authors:** Asish Dasgupta, Kuang-Hueih Chen, Danchen Wu, V. Siddartha Yerramilli, Patricia D.A. Lima, Ashley Y. Martin, Benjamin P. Ott, Jeffrey D. Mewburn, Lian Tian, Ruaa Al-Qazazi, Isaac M. Emon, Pierce Colpman, Lindsay Jefferson, Curtis Noordhof, Oliver Jones, Charles C.T. Hindmarch, Stephen L. Archer

**Author notes:** **Corresponding author: Stephen L. Archer** MD**. FRCP(C)**, **FRSC**, **FAHA**, FACC Scientific Director of TIME (Translational Institute of Medicine) & QCPU (Queen’s CardioPulmonary Unit) Department of Medicine, Queen’s University Elizabeth Smith Distinguished University Professor C. Franklin and Helene K. Bracken Chair Biosciences Complex Room 1520 116 Barrie Street Kingston, Ontario, Canada, K7L 3N6 Preferred. Co-first authors (Asish Dasgupta, Kuang-Hueih Chen).

## Abstract

**Introduction:** Dynamin-related protein 1 (Drp1) activation increases mitochondrial fission and cell cycle progression in hyperproliferative cells, termed *mitotic fission*. However, the diameter of a fission apparatus comprised solely of Drp1 and its binding partners appears insufficient to complete fission. Moreover, the mechanism linking fission to cell cycle progression is unknown, suggesting an additional mediator in the terminal steps of mitotic fission.

**Hypothesis:** The large GTPase, dynamin 2 (DNM2), interacts with Drp1 to complete mitochondrial fission and regulate cell cycle progression.

**Corollary:** DNM2 is epigenetically upregulated in pulmonary arterial smooth muscle cells in pulmonary arterial hypertension (PAH PASMC).

**Methods:** Mitochondrial morphology, protein colocalization and mitochondrial fission sites were assessed using super-resolution microscopy. Protein-protein interaction was confirmed using immunoprecipitation. The role of DNM2’s GTPase domain in mitochondrial targeting was studied by heterologous expression of truncated constructs. Transcriptomic changes from silencing DNM2 in PAH PASMC were measured by RNA-seq. DNM2 expression was quantified in normal and PAH PASMC and lungs from PAH patients and rats with monocrotaline (MCT) and SU5416/hypoxia (Su/Hx)-induced PAH. The effects of manipulating DNM2 on cell proliferation, cell cycle progression and apoptosis were assessed by flow cytometry. siRNA targeting DNM2 was nebulized to rats with MCT-PAH.

**Results:** DNM2 is increased in PAH PASMC in humans and rodents and interacts with Drp1 via its GTPase domain, permitting mitochondrial translocation. Silencing DNM2 in PAH PASMC inhibits fission and slows cell proliferation by causing G1/G0 phase blockade. Augmenting DNM2 in normal PASMC induces fission and accelerates proliferation. siDNM2 downregulates the positive cell cycle regulator, Regulator of Cell Cycle (*RGCC*), in PAH PASMC. Moreover, siRGCC causes G1/G0 cell cycle arrest. miR-124-3p negatively regulates DNM2 and is decreased in PAH PASMC. Augmenting miR-124-3p in PAH PASMC decreases DNM2, inhibits proliferation and induces apoptosis. STAT3 is also negatively regulated by miR-124-3p and siSTAT3 reduces DNM2 and mitochondrial fission in PAH PASMC. Nebulized siDNM2 regresses PAH in MCT-PAH.

**Conclusion:** DNM2’s GTPase domain binds Drp1, mediating mitochondrial translocation and tighter mitochondrial fission. DNM2 is upregulated in PAH by miR-124a-3p deficiency and STAT3 activation. The miR-124a-3p-STAT3-DNM2-Drp1-RGCC pathway underlies accelerated mitotic fission in PAH and offers novel therapeutic targets.

## Introduction

Pulmonary arterial hypertension (PAH) is a lethal syndrome characterized by pulmonary vascular obstruction and stiffening, partially due to a cancer-like hyperproliferation of apoptosis-resistant pulmonary artery smooth muscle cells (PASMC), reviewed in^1^. This *pseudoneoplastic* phenotype is driven, in part, by acquired mitochondrial disorders that promote cell proliferation and impair apoptosis in PAH PASMC, as reviewed in^2^, including **1)** increased uncoupled glycolysis (Warburg metabolism); **2)** disrupted mitochondrial oxygen-sensing, creating a state of pseudohypoxia; **3)** impaired mitochondrial calcium uptake, resulting in reduced intramitochondrial calcium and reduced glucose oxidation; and **4)** an imbalance between fission and fusion, favoring fission. This shift in the fission/fusion ratio regulates cell cycle progression, apoptosis, metabolism, and mitophagy. Approved PAH-targeted therapies are primarily vasodilators and, despite offering symptomatic benefits, functional impairment persists and death from RV failure occurs in 40% of patients within 5-years of diagnosis, as reviewed in^1^. This highlights a need to explore therapeutic targets in other pathways involved in the pathogenesis of PAH, such as PAH’s mitochondria-mediated, *pseudoneoplastic* phenotype.

In rapidly dividing cells there is an obligatory but incompletely understood coordination between mitosis and mitochondrial division. This *mitotic fission* requires activation of dynamin-related protein 1 (Drp1), a large GTPase, and ensures equitable distribution of mitochondria between daughter cells. If mitotic fission is inhibited, by inhibiting fission or enhancing fusion, cell cycle arrest and apoptosis occur, reviewed in^3^. Coordination of fission and mitosis is thought to be mediated, in part, by a shared dependence of both processes on similar kinases, including cyclin B-CDK1^3^, which both activates Drp1 by phosphorylation at serine 616, and triggers mitosis^3^. Upon post-translational activation, Drp1 translocates from the cytosol to the outer mitochondrial membrane (OMM) at a site of narrowing created by contact with the endoplasmic reticulum and actin. There, Drp1 interacts with its binding partners (MiD49, MiD51, MFF and Fis1) and multimerizes, forming a macromolecular fission apparatus that further narrows the mitochondria^4–6^.

In PAH, a mitochondrial-mediated proliferation/apoptosis imbalance, favoring proliferation, occurs in PASMC^7^, endothelial cells^8^ and fibroblasts^9^. Mitochondrial fission is increased in PAH^7^, partly due to upregulation and activation of Drp1. An imbalance between fission and fusion, caused by pathologic Drp1 activation and downregulation of mitofusin-2^10, 11^ favors mitochondrial fission in PAH and cancer cells, reviewed in^12^. These acquired mitochondrial abnormalities are permissive of pathologic rates of cell proliferation and prevent apoptotic removal of abnormal cells. Mitochondrial abnormalities in PAH result both from activation of transcription factors (HIF-1α^13^, NFAT^14^, and STAT3^15^) and epigenetic mechanisms (dysregulation of DNA methyltransferases^13^ and microRNAs (miR)^16^). We successfully targeted increased mitotic fission to regress PAH^7^ and cancer^17, 18^, using small interfering RNA (siRNA) targeting Drp1, small molecule inhibitors of Drp1 GTPase (e.g. mdivi-1^19^ and Drpitor1a, a novel Drp1 inhibitor^20^) or downregulating Drp1’s binding partners, MiD49 and MiD51^18^.

However, the diameter of a fission apparatus comprised solely of Drp1 and its binding partners appears insufficient to complete fission^21, 22^ and the molecular mechanism(s) linking fission to cell cycle progression are incompletely understood. This study explores a putative, terminal step in Drp1-dependent mitochondrial fission which we propose is mediated by the large GTPase, Dynamin 2 (DNM2). We further examine the role of DNM2 in mitochondrial dynamics and cell cycle regulation in normal human PASMC, describe the causes and consequences of DNM2 upregulation in PAH (in patients and multiple rodent models), and exploit DNM2 as a therapeutic target to regress PAH in a rodent model.

## Methods

The data and analytic methods used in this study will be available to other researchers for purposes of reproducing results or replicating the procedure, upon request to the corresponding author.

### Cell culture and reagents

Normal PASMC were either isolated from control subjects or purchased from Lonza (Walkersville, MD, USA) or Cell Applications Inc (San Diego, CA, USA). PAH PASMC were isolated from PAH patients at transplantation or autopsy. All PASMC cell lines were studied within 6 passages. PASMC were cultured in Medium 231 supplemented with SMC Growth Supplement (SMGS, Life Technologies, Carlsbad, CA, USA). Five normal PASMC lines (60% Female, mean age: 65.5 years) and 8 PAH PASMC lines (37.5% Female, mean age: 59.3 years) were used. HEK293A cells were obtained from ATCC (Manassas, Virginia, USA) and cultured in Dulbecco’s modified Eagle’s medium (DMEM) containing 10% FBS, 100 U/ml penicillin, 100 μg/ml streptomycin, and 2 mM glutamine and passaged every 3 days. See supplemental materials for information on culturing Drp1 knockout (KO) mouse embryonic fibroblasts (MEFS).

### siRNA and primers

See supplemental materials.

### Immunohistochemistry

DNM2 expression was assessed using immunohistochemistry in human lungs (normal, n=5, PAH, n=5, demographics in supplementary Table 1) and rodent lungs (Su/Hx rat model: n=4/group; Su/Hx mouse model; n=4/group, MCT rat model, n=5/group), as described^23^. DNM2 expression was quantified by the signal intensity of brown colour in the medial layer of small PAs in the lungs using ImageJ (NIH, Bethesda, MD, USA).

### Immunofluorescence

Immunofluorescence staining of the formalin-fixed paraffin-embedded rat lung tissues was performed as described^24^. See supplemental material.

### Western blotting

See supplemental materials.

### Immunoprecipitation

Lysis buffer (Cell Signaling Technologies, Beverly, MA, USA) supplemented with a protease and phosphatase inhibitor cocktail (Thermo Fisher Scientific, Waltham, MA, USA) was used to lyse cell pellets. Cell lysates were incubated with anti-Drp1 antibody (2 µg, Becton Drive, NJ, US) at 4°C overnight. For detailed methods see supplemental material.

### siRNA and plasmid transfection

PAH PASMC were grown to 60-80% confluence and then transfected with 25 picomoles of siRNA, as described^23^. The knockdown efficiency was assessed after 48 hours using qRT-PCR (Bio-Rad, Hercules, CA, USA) and immunoblotting. For plasmid overexpression, the cells were transfected with 2.5 μg of the specified plasmid, as described^23^, also see supplemental material.

### Cell cycle analysis

The DNA content of PAH PASMCs was analyzed with propidium iodide (PI) 72 hours post-transfection using flow cytometry (Sony SH800S, San Jose, CA, USA). See supplemental material.

### Cell proliferation assay

Cell proliferation was assessed 72-hours following transfection with either siRNA or plasmids, using the Click-iT EdU kit following manufacturer’s instructions (Life Technologies, Carlsbad, CA, USA).

### Apoptosis Assay

Apoptosis was determined in PAH PASMC 72 hours following transfection by the Annexin V+/PI method, using flow cytometry, as described^18^ and detailed in supplemental material.

### Confocal and Stimulated Emission Depletion (STED) Microscopy of Live cells

Live cells were transfected with specified siRNAs or miRs and imaged with a Leica SP8 laser scanning confocal microscope (Leica Microsystems, Wetzlar, Germany), as previously described^23^. For detailed methods, please see the supplemental material.

### Confocal and STED Imaging of Fixed Cells

For detailed methods see supplemental material.

### Image Analysis

Image analysis was performed using ImageJ and Leica Application Suite X (LAS X) software. The Pearson Correlation Coefficient was quantified using ImageJ and LAS X software.

### Quantification of mitochondrial morphology and networking

Mitochondrial fission was quantified using three techniques. 1) Mitochondrial morphology was quantified by measuring mitochondrial fragmentation count (MFC) with reduced MFC indicating a more fused mitochondrial network. 2) Machine learning algorithm that independently classified mitochondria as punctate, intermediate or filamentous, as described^17, 23^. 3) Mitochondrial networking analyses in which cells are transfected with a mitochondrial-targeted, photoactivatable, green fluorescent protein (mito-PA-GFP) to track the diffusion of GFP along the mitochondrial lumen using a metric called the mitochondrial networking factor (MNF) which increases in proportion to fusion, as described ^17, 23^.

### miR-124-3p and STAT3 binding luciferase reporter assays

The binding of miR-124a-3p to the 3’-UTR of DNM2’s mRNA was validated using a luciferase binding assay, as previously described^18, 23^. The binding of STAT3 to DNM2’s promoter region was validated using a promoter binding assay, as described in the supplemental material.

### Ethical Approval

The animal experiment protocol was approved by the University Animal Care Committee of Queen’s University (protocol# 2021-2112). The study of human cells and tissues was approved by the Queen’s University Health Sciences and Affiliated Teaching Hospitals Research Ethics Board (#6014017).

### MCT-PAH model

PAH was induced in adult, 200-250g, male and female, Sprague-Dawley rats (Charles River, NY, USA) by a single subcutaneous injection (sc) of monocrotaline (60mg/kg), as described^25^. PBS was used as a control vehicle.

### *In vivo* siRNA treatment

Briefly, 1 nanomole of siDNM2 or ctrl-siRNA, modified with orthothioates to increase stability, (Integrated DNA Technology, Coralville, IA, USA) was administered beginning on day 15, after PAH had been detected and was repeated on days 18, 21, and 24 post-MCT injection. The in vivo siRNA was nebulized (50μl saline using an aerosol nebulizer, Kent Scientific cat #neb-1200) to anesthetized rats for lung-specific delivery.

### Pulmonary Hemodynamics

Pulmonary hemodynamics were assessed by right heart catheterization (RHC) on day 25 post-MCT injection via a closed-chest approach. Right ventricular systolic pressure (RVSP) and cardiac output (CO) were determined as described^26^. Subsequently, left-ventricular end-diastolic pressure (LVEDP) was measured via a closed-chest, retrograde catheterization via the right carotid artery. Cardiac index (CI), mean pulmonary artery pressure (mPAP) and pulmonary vascular resistance index (PVRI) were calculated, as described in the supplemental material.

### Rat SU5416/hypoxia rat model

SU5416/hypoxia PAH model (Su/Hx) was created in Sprague-Dawley rats of both sexes by injection of SU5416 (20mg/kg sc). Rats were housed in a hypoxic chamber with 10% oxygen for 3 weeks and then returned to normoxia for 3 weeks before sacrifice and PASMC isolation, as described^10^.

### Mouse SU5416/hypoxia model

Mice from both sexes were placed in a normobaric chamber with an incremental decrease of oxygen levels to 10% over 3 days. They were maintained for 3 weeks at 10% FiO2 without returning to normoxia. Animals were given SU5416 (20mg/kg sc) on days 0, 7 and 14 of hypoxia. Echocardiography was performed on day 21 with RHC and tissue collection on day 22. The development of PAH was confirmed as shown in Supplemental Fig. 1A-D.

### Statistical analysis

Data are presented as the mean ± standard error of the mean (SEM). Data were tested for normality using Kolmogorov-Smirnov Test and then a Student t-test or Mann-Whitney U test (unpaired or paired) was used to compare the mean or median value between two groups, as appropriate. Intergroup differences of 3 or more groups were assessed using an ANOVA, with Šídák’s multiple comparisons test. Two-way ANOVA with the Holm-Šídák test post hoc analysis was used for machine learning-based mitochondrial morphology analysis. *P* < 0.05 was considered statistically significant. Analyses were performed using GraphPad-Prism 10 software (San Diego, CA, USA).

The RNA sequencing analysis was conducted with the DESeq2^27^ package (v1.44) in R (v4.4.0), which uses a negative binomial generalized linear model for differential expression analysis of RNA-seq count data.Best laboratory practices were implemented by use of animals and patients of both sexes, randomization of groups, using of a blinding system for data acquisition, and use of adequate sample size, as described^28^.

## Results

### DNM2 expression in PASMC is increased in human and rodent PAH

Increased DNM2 expression occurred in PASMC from PAH patients (Fig. 1A-B). DNM2 is also increased in the media of pulmonary arterioles (PA) in PAH patients, in rats with MCT-PAH or Su/Hx-PAH, and in mice with Su/Hx-PAH (Fig. 1C-F). Conversely, the other two members of the dynamin family, DNM1 and DNM3, were not upregulated in human PAH PASMC (Supplementary Fig. 2A-B).

**Figure 1:**
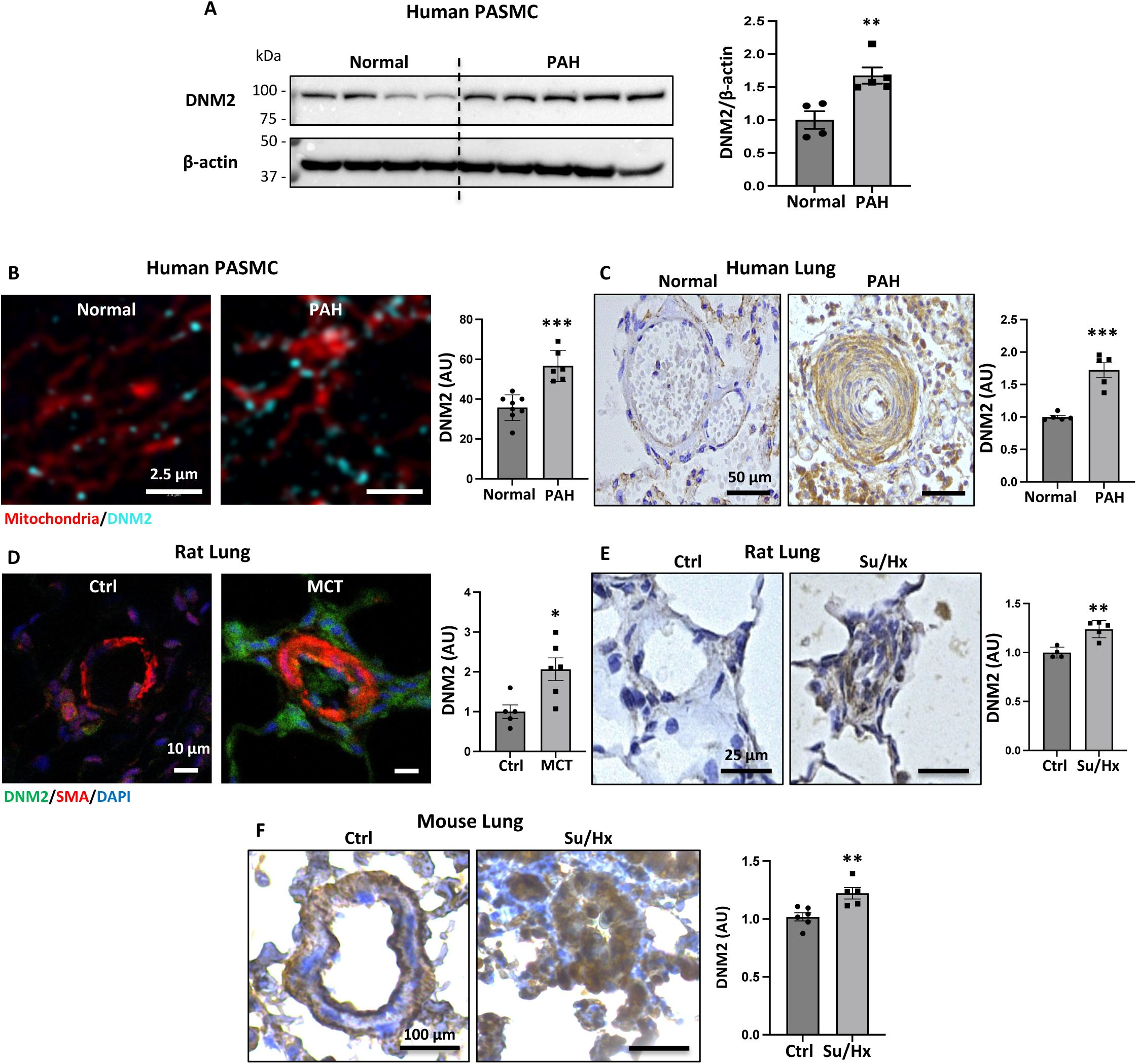
A) Representative immunoblot and densitometry showing increased expression of DNM2 in human PAH PASMC (n=5) vs normal human PASMC (n=4). β-actin was used as the loading control (\*\**P* < 0.01). **B) Representative STED super-resolution confocal microscopy images demonstrating increased expression of DNM2 in PAH PASMC.** Staining used in the images to create colors: mitochondria (red, MitoTracker^TM^ Deep Red) and DNM2 (cyan) in normal and PAH PASMC. Scale bar: 2.5 μm. (\*\*\**P* < 0.001). **C) Human lungs demonstrate increased expression of DNM2 in the media of small PAs in PAH vs control subjects.** Representative images and quantification of immunohistochemistry demonstrating increased expression of DNM2 (brown) in the media of small PAs from human PAH lungs vs control lungs (n=5-7 PAs from each subject, 5 subjects per group, < 100 μm in diameter). Scale bar: 50 μm. (\*\*\**P* < 0.001). **D) Immunofluorescence showing increased expression of DNM2 (green) in the media of distal PAs from MCT-PAH vs control rats.** Representative images and quantification of immunofluorescence demonstrating DNM2 (green) in the media (red) of small PAs from control vs MCT-PAH lungs (n=5-10 PAs from each rat, 5 rats per group, < 100 μm in diameter). Scale bar: 10 μm. (\**P* < 0.05). **E) Immunohistochemistry showing increased DNM2 (brown) in the media of small PAs in Su/Hx rats.** Representative images and quantification of immunohistochemistry demonstrating increased expression of DNM2 (brown) in the media of PAs from Su/Hx-PAH lungs vs control lungs (n=6-9 PAs from 4 controls and 5 Su/Hx rats, < 100 μm in diameter). Scale bar: 25 μm. (\*\**P* < 0.01). **F) Immunohistochemistry showing increased DNM2 (brown) in the media of small PAs of Su/Hx mice.** Representative images and quantification of immunohistochemistry demonstrating increased expression of DNM2 (brown) in the media of PAs from Su/Hx-PAH lungs vs normoxia (Nx) control lungs (n=4-5 PAs from 3 control mice and 4 Su/Hx mice, < 100 μm in diameter). Scale bar: 100 μm. (\*\**P* < 0.01).

### Silencing DNM2 inhibits mitochondrial fission in PAH PASMC whilst DNM2 overexpression causes fission in normal PASMC

siDNM2 reduced DNM2 mRNA and protein expression by 84% and 71%, respectively (Supplementary Fig. 3A-B). siDNM2 in PAH PASMC inhibits mitochondrial fission, creating a filamentous network, reducing MFC^17^ and increasing filamentous mitochondria, as detected by a bias-free machine learning algorithm^23^ (Fig. 2A). siDNM2 knockdown also significantly increased MNF^17^, reflecting increased diffusion of mito-PA-GFP through the mitochondrial network (Fig 2B).

**Figure 2.**
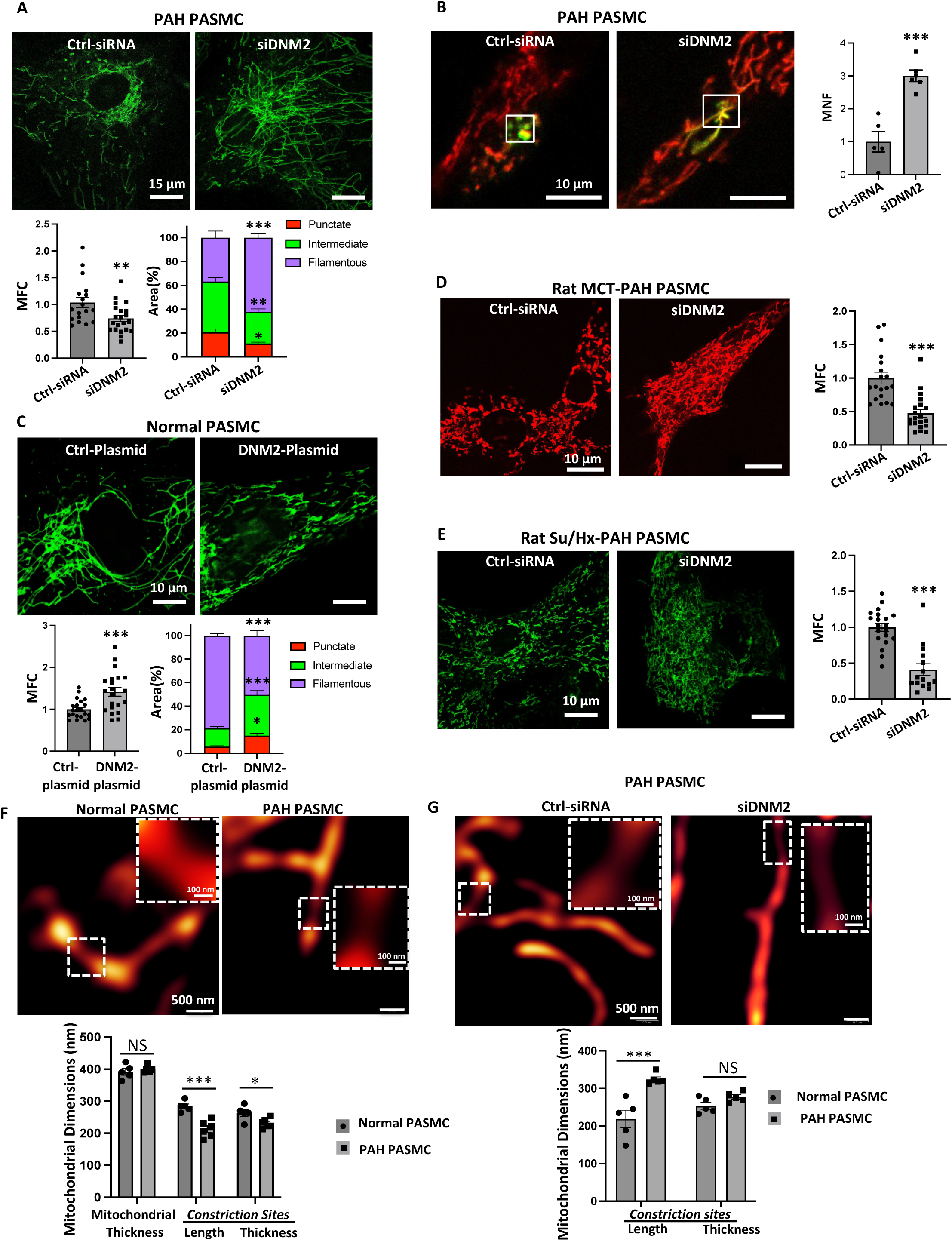
DNM2 regulates mitochondrial fission in human PASMC. A) Silencing DNM2 causes mitochondrial fusion in PAH PASMC. Representative images of mitochondrial networks of PAH PASMC. PAH PASMC were transfected with the specified siRNA and infected with Adv-mNeon Green and imaged after 48h following infection. Mitochondrial fragmentation was quantified by mitochondrial fragmentation count (MFC), and machine learning was used to calculate the percentage area of punctate, intermediate and filamentous mitochondria of each image. Scale bar: 15 μm. (\**P* < 0.05, \*\**P* < 0.01, \*\*\**P* < 0.001; n=18-21 cells/group). **B) Silencing of DNM2 increases mitochondrial networking factor (MNF).** Representative images of mitochondrial network in cell transfected with mito-PA-GFP, quantified by determining MNF. MNF is increased in PAH PASMC following silencing DNM2, indicating restoration of fusion (n=5 cells/group). Scale bar: 10 μm. (\*\*\**P* < 0.001). **C) Augmenting DNM2 in normal human PASMC induces mitochondrial fission.** Representative images of mitochondrial networks of normal human PASMC transfected with the specified plasmid and infected Adv-mNeon Green and imaged after 48h following infection. Mitochondrial fragmentation was quantified by MFC and a machine learning algorithm (n=21 cells/group). Scale bar: 10μm. (\**P* < 0.05, \*\*\**P* < 0.001). **D) Silencing DNM2 causes mitochondrial fusion in rat MCT-PAH PASMC.** Representative images of mitochondrial networks of rat MCT-PAH PASMC. Rat MCT-PAH PASMC were transfected with the specified siRNA and imaged after 48h following loading the cells with TMRM. Mitochondrial fragmentation was quantified by MFC (n=19-20 cells/group). Scale bar: 10 μm. (\*\*\**P* < 0.001). **E) Silencing DNM2 causes mitochondrial fusion in rat Su/Hx-PAH PASMC.** Representative images of mitochondrial networks of Su/Hx-PAH PASMC transfected with the specified siRNA, stained with MitoTracker^TM^ Green and imaged after 48h following infection. Mitochondrial fragmentation was quantified by MFC (n=15-20 cells/group). Scale bar: 10μm. (\*\*\**P* < 0.001). **F-G) DNM2 regulates the dimensions of the mitochondrial constriction sites in human PASMC**. **F) Dimensions of mitochondrial constriction sites are decreased in PAH PASMC.** Representative confocal microscopic images and quantification of the dimensions of mitochondrial constriction sites in normal vs PAH PASMC. Mitochondria were stained using MitoTracker^TM^ Deep Red (n=5-6 cells/group, 44-79 measurements/cell) for length and (n=5-6 cells/group, 43-74 measurements/cell) for thickness of the mitochondrial constriction sites. Scale bar = 500 nm. (\**P* < 0.05, \*\*\**P* < 0.001, NS, not significant). **G) Silencing DNM2 increases length of mitochondrial constriction sites in PAH PASMC.** Representative STED images and quantification of the dimensions of the mitochondrial constriction in PAH PASMC transfected with specified siRNA for 48h and stained with MitoTracker^TM^ Deep Red (n=5 cells/group, 22 measurements/cells of length and thickness of mitochondrial constriction sites). Scale bar: 500 nm. (\*\*\**P* < 0.001, NS: not significant).

Conversely, overexpressing DNM2 in normal PASMC increased mitochondrial fission, recapitulating a PAH phenotype (Fig. 2C). Similar to human PAH PASMC, mitochondria in rat MCT-PAH PASMC and Su/Hx-PAH PASMC (studied within passage 1-2) exhibited increased mitochondrial fission^23^. Likewise, siDNM2 inhibited mitochondrial fission and increased fusion of the mitochondrial network in rat MCT-PAH PASMC and Su/Hx-PAH PASMC (Fig. 2D-E).

Silencing Drp1 does not alter expression of DNM2 nor vice versa (Supplementary Fig. 4A), indicating the effects of siDNM2 are not indirectly mediated by Drp1 suppression. Moreover, silencing DNM2 in Drp1-silenced PAH PASMC did not further inhibit mitochondrial fission, indicating that DNM2’s pro-fission ability is Drp1-dependent (Supplementary Fig. 4B). Silencing DNM2 in PAH PASMC also did not alter the expression of other fission and fusion mediators, indicating that siDNM2’s effect on PASMC is due to inhibition of DNM2-mediated mitochondrial fission, rather than off-target effects (Supplementary Fig. 5A-D). Furthermore, the mitochondria of PAH PASMC displayed an increased invagination at evolving mitochondrial fission sites as evidenced by a decrease in length and residual thickness of these mitochondrial constriction sites as measured by quantifying the fluorescence intensity of two mitochondrial vital dyes namely MitoTracker Deep Red and MitoTracker Green using STED super resolution, microscopy (Fig. 2F and Supplementary Fig. 5E). Neither MitoTracker Deep Red nor MitoTracker Green are sensitive to transient changes in mitochondrial membrane potential, thus narrowing in mitochondrial constriction sites are not artifacts of transient mitochondrial membrane depolarization, as could occur with TMRM. Further demonstrating DNM2’s role in creating short and tight areas of constriction, silencing DNM2 in PAH PASMC produced significant elongation of the mitochondrial constrictions sites (Fig. 2G).

### DNM2-induced mitochondrial fission is Drp1-dependent and requires an intact DNM2 GTPase domain

STED microscopy shows that there is an increase in the co-localization between Drp1 and DNM2 in PAH PASMC (Fig. 3A-i-ii), evident as a reduced Drp1-DNM2 intermolecular distance (Fig. 3A-iii). Interestingly, increased Drp1-DNM2 co-localization at the mitochondrial constriction sites is associated with elevated DNM2, rather than Drp1 expression (Fig. 3A-iv). The proximity of DNM2 and Drp1 on the mitochondrial membrane were further quantified by placing a linear region of interest (ROI) along the long axis of the mitochondrion. The proximity between Drp1 and DNM2 molecules is decreased in PAH versus control PASMC (Fig. 3B). Consistent with this STED microscopy finding, co-immunoprecipitation studies confirmed the direct interaction between DNM2 and Drp1 (Fig. 3C-D).

**Figure 3.**
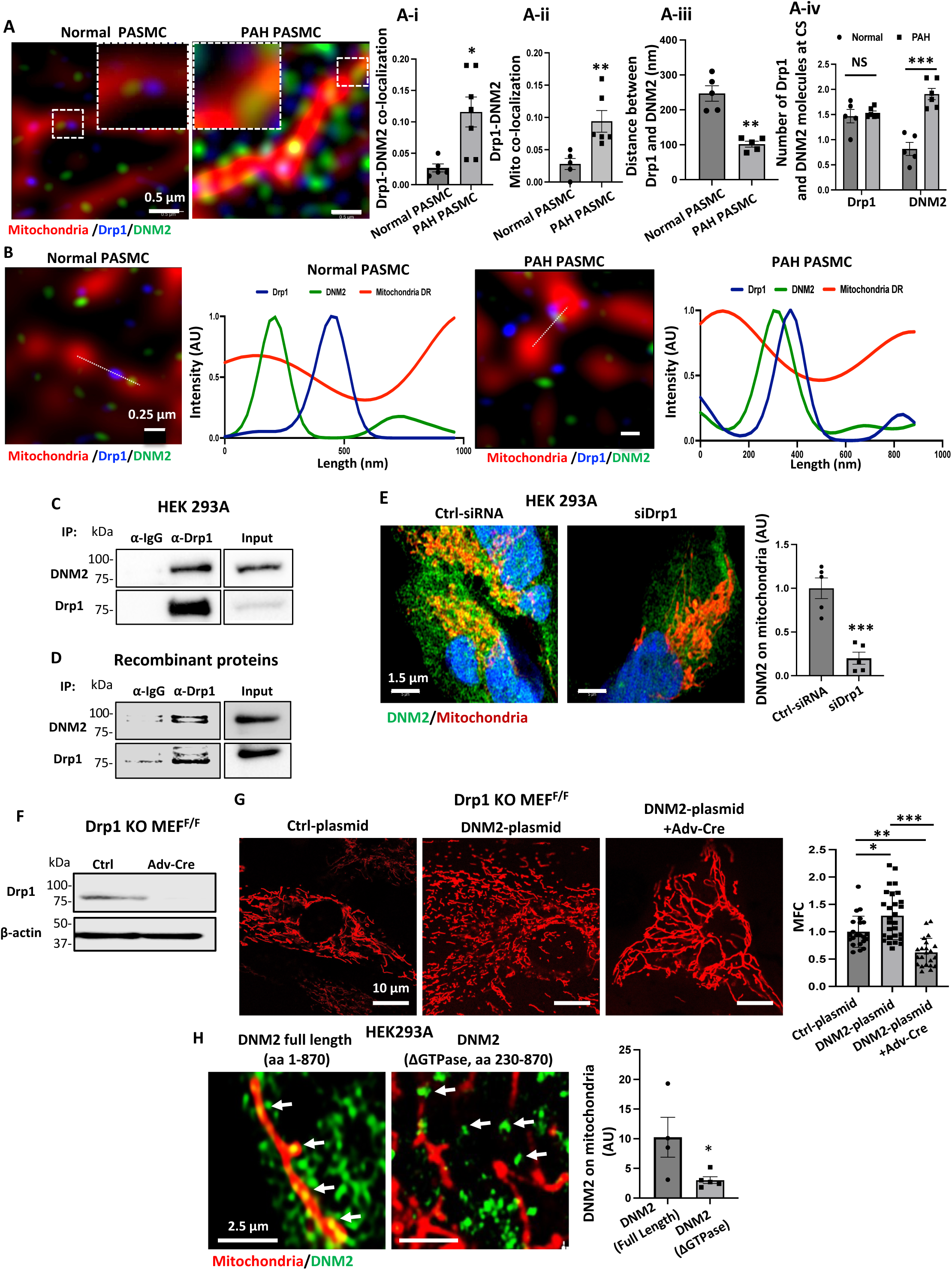
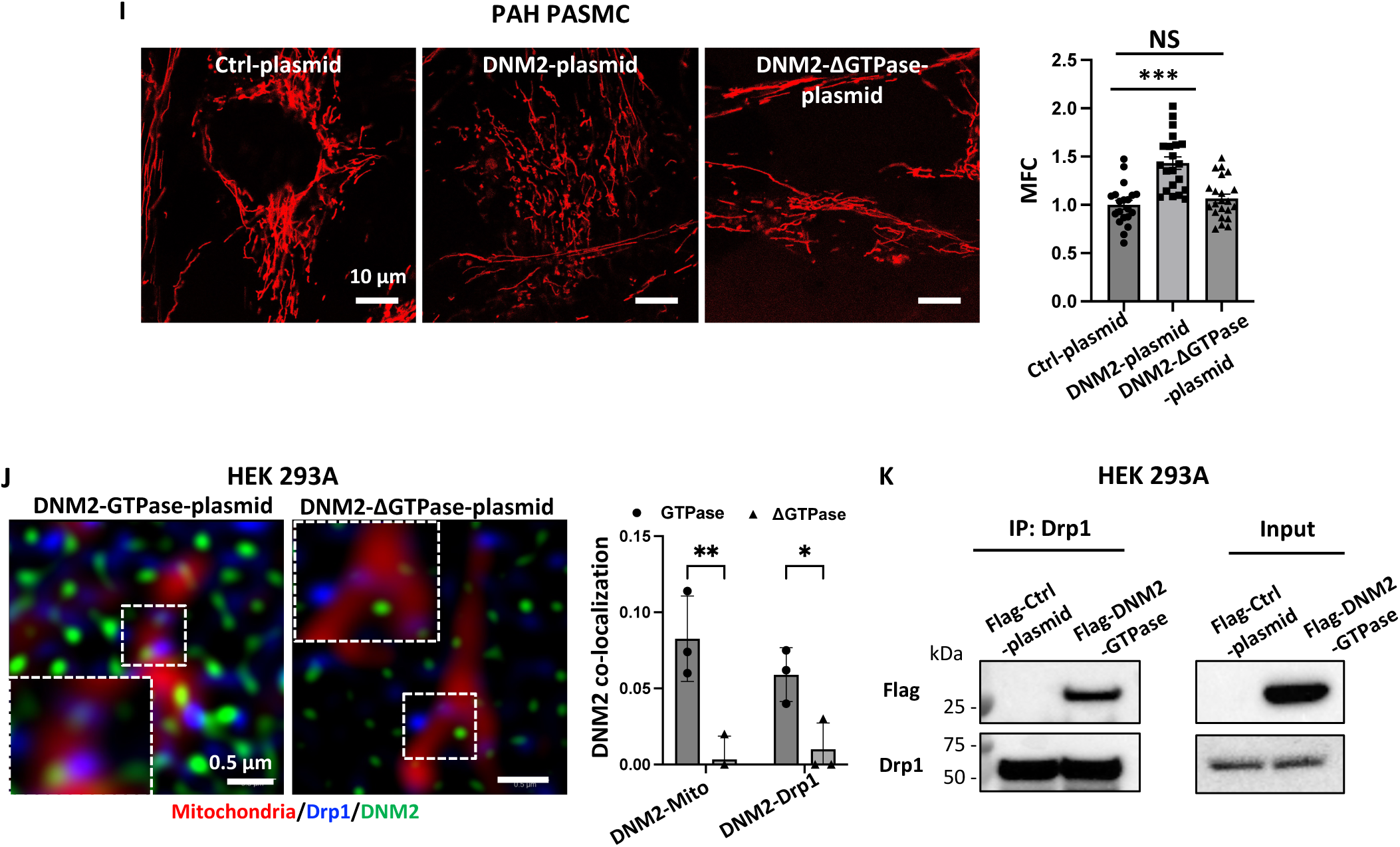
DNM2-induced mitochondrial fission is Drp1-dependent. **A-B) DNM2 and Drp1 co-localize on mitochondria at the mitochondrial constriction sites (CS) of pulmonary artery smooth muscle cells.** **A) Representative STED images and quantification showing colocalization between Drp1 (blue), DNM2 (green) molecules on mitochondria (red) in normal and PAH PASMC**. A-i) Whole cell colocalization was quantified using Pearson’s coefficient (n=5-7 cells/group). A-ii) Mitochondrial co-localization was quantified using Pearson’s coefficient (n=5-6 cells/group). A-iii) Quantification of distance between individual Drp1 and DNM2 molecules on mitochondria (n=5 cells/group). A-iv) Quantification of the number of individual Drp1 and DNM2 molecules on the mitochondrial CS (n=5-6 cells/group). Scale Bar: 0.5 μm. (\**P* < 0.05, \*\**P* < 0.01, \*\*\**P* < 0.001, NS, not significant). **B) Increased co-localization between Drp1 and DNM2 at the mitochondrial constriction sites.** ROIs drawn along mitochondrial constriction sites (CS) on a normal PASMC (left) and PAH PASMC (right). Plot profile of averaged fluorescent intensities of Drp1, DNM2 and MitoTracker^TM^ Deep Red stain along the ROIs (n=39-42 points along the ROIs/group). Scale bar: 0.25 μm. **C-D) Drp1 and DNM2 interact with each other**. **C)** Co-immunoprecipitation reaction performed with cell lysates (1000 μg) from HEK293A. Immunoprecipitation reaction performed with anti-Drp1 antibody and immunoblotted with anti-DNM2 antibody (top panel) and total Drp1 antibody (bottom panel). Total cell extracts (100 μg) were used as positive control for the expression of DNM2 and Drp1. **D)** Co-immunoprecipitation reaction performed with 300 ng of DNM2 and 300 ng of Drp1 recombinant proteins. The immunoprecipitation reaction was performed with anti-Drp1 antibody and immunoblotted with anti-DNM2 antibody (top panel) and total anti-Drp1 antibody (bottom panel). 100 ng of DNM2 and Drp1 recombinant proteins were used as positive control for the presence of DNM2 and Drp1. **E) DNM2 mediated mitochondrial fission is Drp1 dependent.** DNM2 (green) fails to localize to mitochondria (red) when Drp1 is eliminated (i.e. less yellow following siDrp1). Representative confocal images and quantification of mitochondrial localization of DNM2 in HEK293A cells transfected with GFP-DNM2 and co-transfected with either ctrl-siRNA or siDrp1. 48 h following transfection the mitochondria were stained with MitoTracker^TM^ Deep Red (n=5 cells/group). Scale bar=1.5 μm. (\*\*\**P* < 0.001). **F-G) Augmenting DNM2 failed to induce mitochondrial fission in Drp1 conditional knockout MEFs**. **F)** Representative immunoblot showing effective knockout of Drp1 in Drp1 conditional knockout MEFs following infection with Adv-Cre for 48 h. **G)** Representative images of mitochondrial networks and quantification of mitochondrial fragmentation in Drp1 conditional knockout MEFs transfected with the specified plasmids and infected with Adv-Cre to knockout Drp1 and loaded with TMRM 48 h post infection (n=22-28 cells/group). Scale bar: 10 μm. (\**P* < 0.05, \*\**P* < 0.01, \*\*\**P* < 0.001). **H-K) DNM2’s GTPase domain is critical for mitochondrial localization and fission via interaction with Drp1**. **H) DNM2’s GTPase domain is critical for mitochondrial localization.** Representative STED microscopic images of mitochondria of HEK293A cells show full length DNM2 (green) is localized on the red mitochondria stained with MitoTracker^TM^ Deep Red (yellow dots) whilst a deleted GTPase domain, GFP fusion construct (ΔGTPase, aa 230-870) failed to localize to the mitochondrial OMM (n=4-5 cells/group). Scale bar=2.5 μm. (\**P* < 0.05). **I) DNM2-ΔGTPase failed to induce mitochondrial fission in PAH PASMC.** Representative images of mitochondrial network and quantification of mitochondrial fragmentation in PAH PASMC transfected with specified plasmids and loaded with TMRM, 48 h post-transfection (n=22 cells/group). Scale bar=10 μm. (\*\*\**P* < 0.001, NS: not significant). **J) DNM2 GTPase co-localize with Drp1.** STED super-resolution images of cells transfected with GFP tagged DNM2-GTPase or DNM2-ΔGTPase constructs (green), and subsequently stained for Drp1 (using anti-Drp1 antibody, blue) and for mitochondria (using MitoTracker^TM^ Deep Red, red). Co-localization between DNM2 and Drp1 on mitochondria was quantified using Pearson’s coefficient (n=5 cells/group). Scale bar=0.5 μm. (\**P* < 0.05, \*\**P* < 0.01). **K) The GTPase domain of DNM2 interacts with Drp1.** HEK293A cells were transfected with Flag-Ctrl or Flag-DNM2-GTPase domain plasmids for 48 h. Co-immunoprecipitation was performed with cell lysates (2000 μg) with anti-Drp1 antibody and immunoblotted with anti-Flag antibody (top panel) and total Drp1 antibody (bottom panel). Total cell extracts (100 μg) were used as positive control for the expression of DNM2-GTPase and Drp1.

In addition, silencing Drp1 inhibited the translocation of DNM2 to mitochondria (Fig. 3E), consistent with Drp1 being involved in DNM2 trafficking, which is relevant since DNM2 lacks a mitochondrial targeting sequence. Further supporting an obligatory role of Drp1 in DNM2-mediated mitochondrial fission, augmenting DNM2 increased mitochondrial fission in conditional Drp1-knockout MEFs (without Adv-Cre) in the control state; however, DNM2-induced fission was eliminated in this cell line when Drp1 was deleted using adenovirus-delivered Cre (Fig. 3F-G). In addition, a DNM2 construct lacking the GTPase domain (ΔGTPase) fails to localize to mitochondria or induce mitochondrial fission (Fig. 3H-I). Conversely, a construct containing only DNM2’s GTPase domain localized on mitochondria and interacted with Drp1 (Fig. 3J-K and Supplementary Fig. 6A-B). Furthermore, pharmacological inhibition of DNM2’s GTPase activity inhibited DNM2’s mitochondrial localization (Supplementary Fig. 7). These three findings in aggregate demonstrate that the GTPase domain of DNM2 is critical for both its Drp1-dependent mitochondrial localization and its pro-fission properties and support an essential role for DNM2 in fission.

### DNM2 regulates cell proliferation, cell cycle and apoptosis in PASMC

Silencing DNM2 inhibited proliferation of human PAH PASMC (Fig. 4A), which was accompanied by G1/G0 phase cell cycle arrest (Fig. 4B). Furthermore, siDNM2 downregulated the expression of the cell cycle protein, CDK4, and its regulator cyclin D1, and upregulated the inhibitor of CDK4, p27^Kip1^ (Fig. 4C-E), identifying the potential inhibitory mechanism by which reducing DNM2 expression effects slowing of cell proliferation.

**Figure 4.**
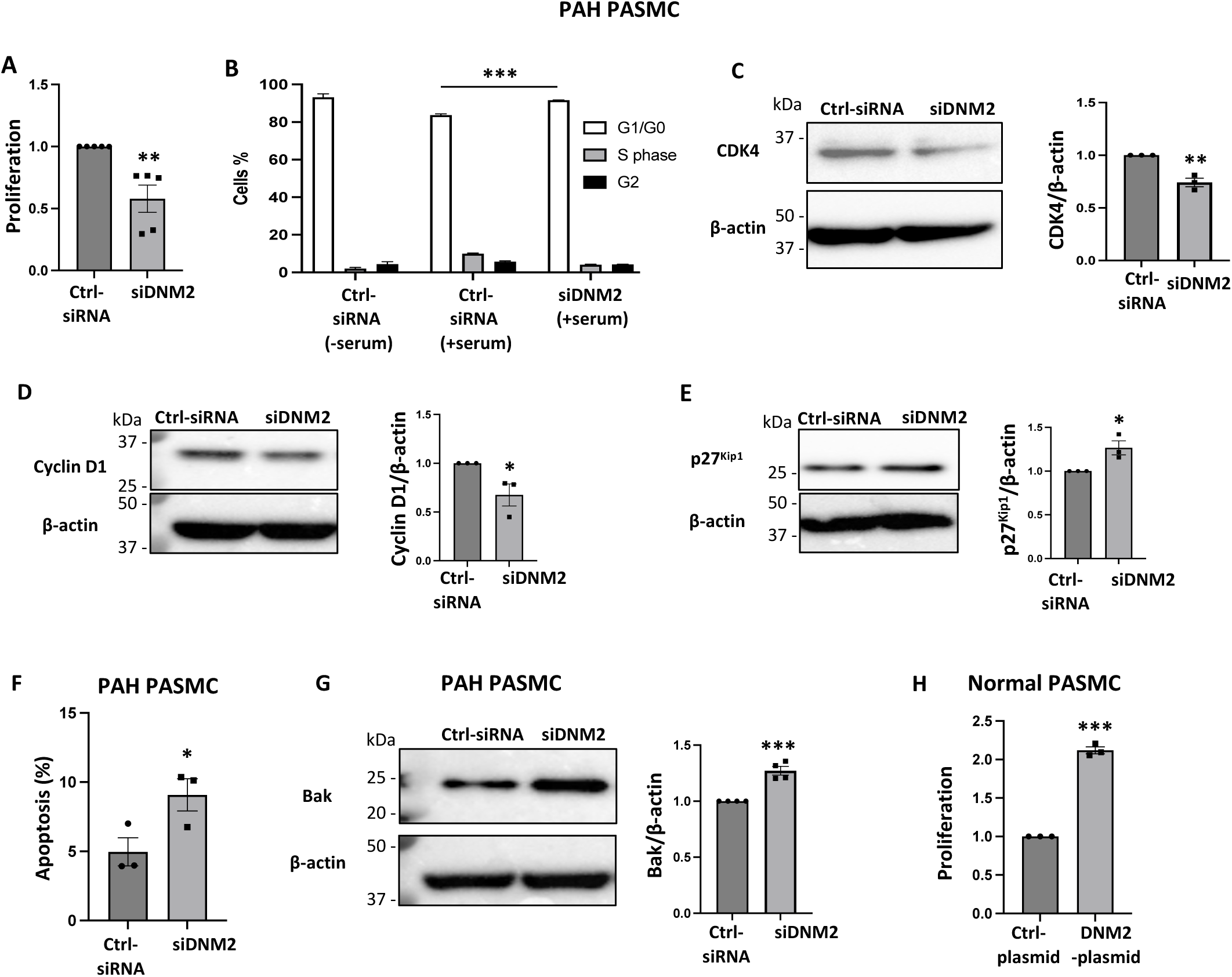
DNM2 regulates proliferation and apoptosis in PASMC. A) Silencing DNM2 inhibits proliferation of PAH PASMC. Cell proliferation was analyzed by EdU incorporation assay 72 h post-transfection (n=5 PAH cell lines/group) (\*\**P* < 0.01). **B) Silencing DNM2 induces cell cycle arrest in the G1/G0 phase.** PAH PASMC was transfected with siDNM2 for 24 h, serum starved for 24 h followed by serum stimulation for 24 h. Cell cycle analyses were performed by flow cytometry following propidium iodide (PI) staining (n=3 technical repeats) (\*\*\**P* < 0.001). **C-E) Silencing DNM2 inhibits the expression of cell cycle proteins CDK4 and Cyclin D1 while increasing the expression of the cell cycle inhibitor, p27^Kip1^.** Representative immunoblots and densitometries of the expression of **C)** CDK4, **D)** Cyclin D1 and **E)** p27^Kip1^. PAH PASMC were transfected with specified siRNA. Cells were harvested for immunoblot analyses 48 h following transfection. β-actin was used as the loading control (n=3 PAH cell lines/group) (\**P* < 0.05, \*\**P* < 0.01). **F-G) Silencing of DNM2 induces apoptosis in PAH PASMC and upregulates the apoptotic mediator BAK. F)** PAH PASMC were transfected with specified siRNA for 72 h. The cells were labeled with Annexin V^FITC^ and PI and assessed by flow cytometry analyses (n=3 PAH cell lines/group). (\**P* < 0.05). **G)** Representative image of immunoblot and densitometry of the expression of Bak. PAH PASMC were transfected with specified siRNA for 48 h. Cells were harvested for immunoblot analyses 48 h following transfection. β-actin was used as the loading control (n=4 PAH cell lines/group) (\*\*\**P* < 0.001). **H) Augmenting DNM2 induces proliferation of normal human PASMC.** Normal human PASMC were transfected with specified plasmids. Cell proliferation was analyzed by EdU incorporation assay 72h post-transfection (results of one representative cell line from n=3 normal PASMC cell lines) (\*\*\**P* < 0.001).

In addition, silencing DNM2 increased unstimulated apoptosis in PAH PASMC (Fig. 4F) and, consistent with this, increased the expression of the apoptotic mediator Bak (Fig. 4G). Conversely, augmenting DNM2 expression in normal PASMC induced proliferation, recapitulating a PAH phenotype (Fig. 4H). Similar to human PAH PASMC, MCT and Su/Hx rat PASMC maintained a hyperproliferative phenotype in culture (both within passage 1-2) which was reversed by transfection of siDNM2 (Supplementary Figs. 8A-B). Silencing DNM1 also inhibits proliferation of human PAH PASMC whereas silencing DNM3 did not (Supplementary Figs. 9A-B); however, since DNM1’s expression is unchanged in PAH PASMC, it was excluded from further study.

### DNM2 regulates cell cycle through Regulator of cell cycle (*RGCC*)

We next investigated the mechanistic pathway that connects DNM2 to the cell cycle. We treated human PAH PASMC cell lines with siDNM2 and used RNA sequencing (RNA-seq) to identify potential DNM2-regulated cell cycle mediators. 140 genes passed the filtering threshold of an adjusted p-value of 0.1, indicating differential expression after silencing DNM2 (Fig. 5A). Of all the down-regulated genes, DNM2 has the lowest log_2_ (fold-change) of -1.388 and the highest -log_10_ (adjusted p-value) of 9.102, which indicates successful silencing of DNM2. The other most down-regulated genes are *RGCC, TICAM1, RASSF3,* and *MRPL39* (Table 2). *RGCC*, the most downregulated gene, following DNM2 silencing, is a known regulator of cell cycle progression^29,30^. Gene Ontology (GO) analysis of the siDNM2-regulated genes revealed that *RGCC* is enriched in most of the GO terms consisting of “cell cycle” (Fig. 5B and Supplementary Table 3). Therefore, we selected RGCC for further investigation.

**Figure 5.**
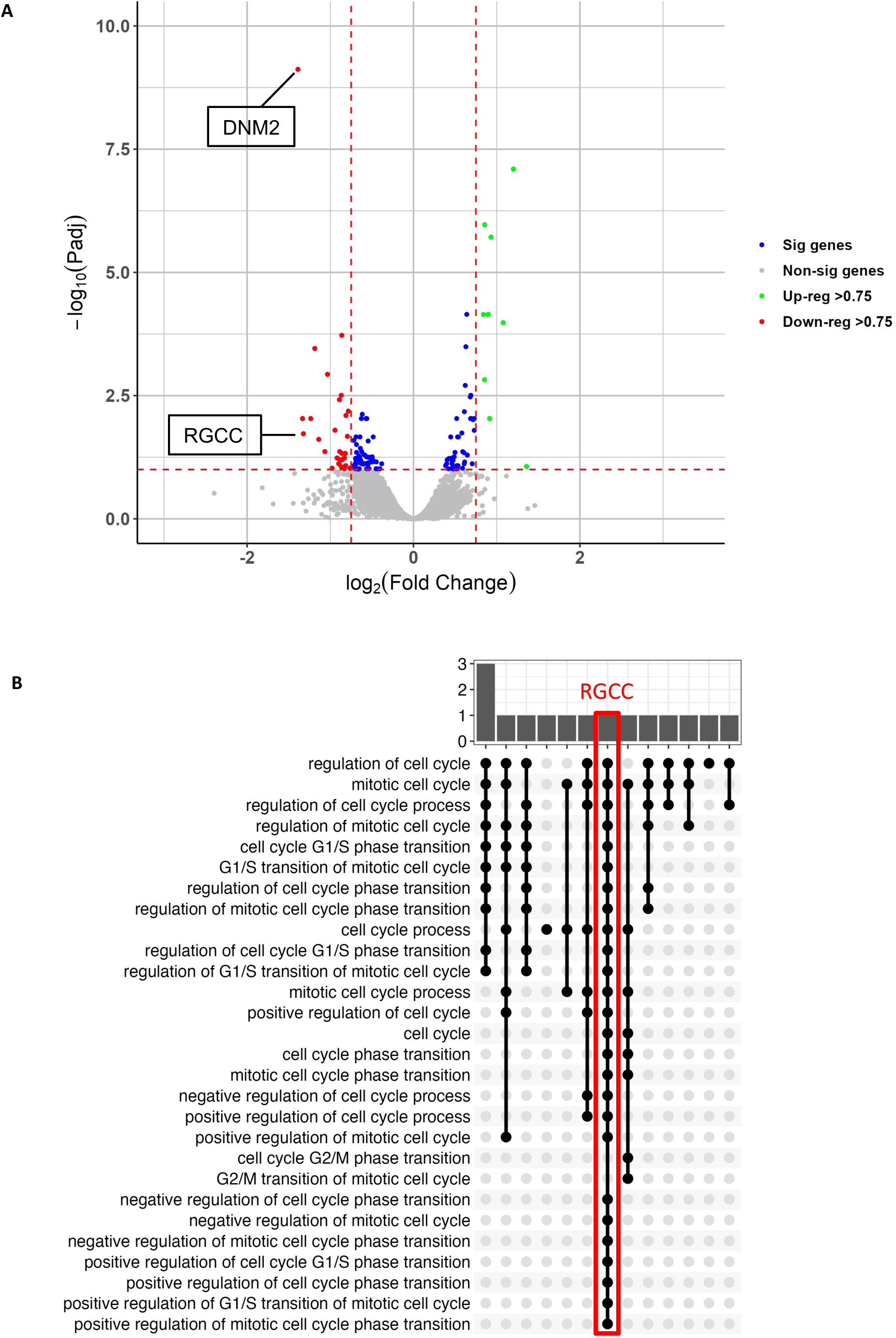
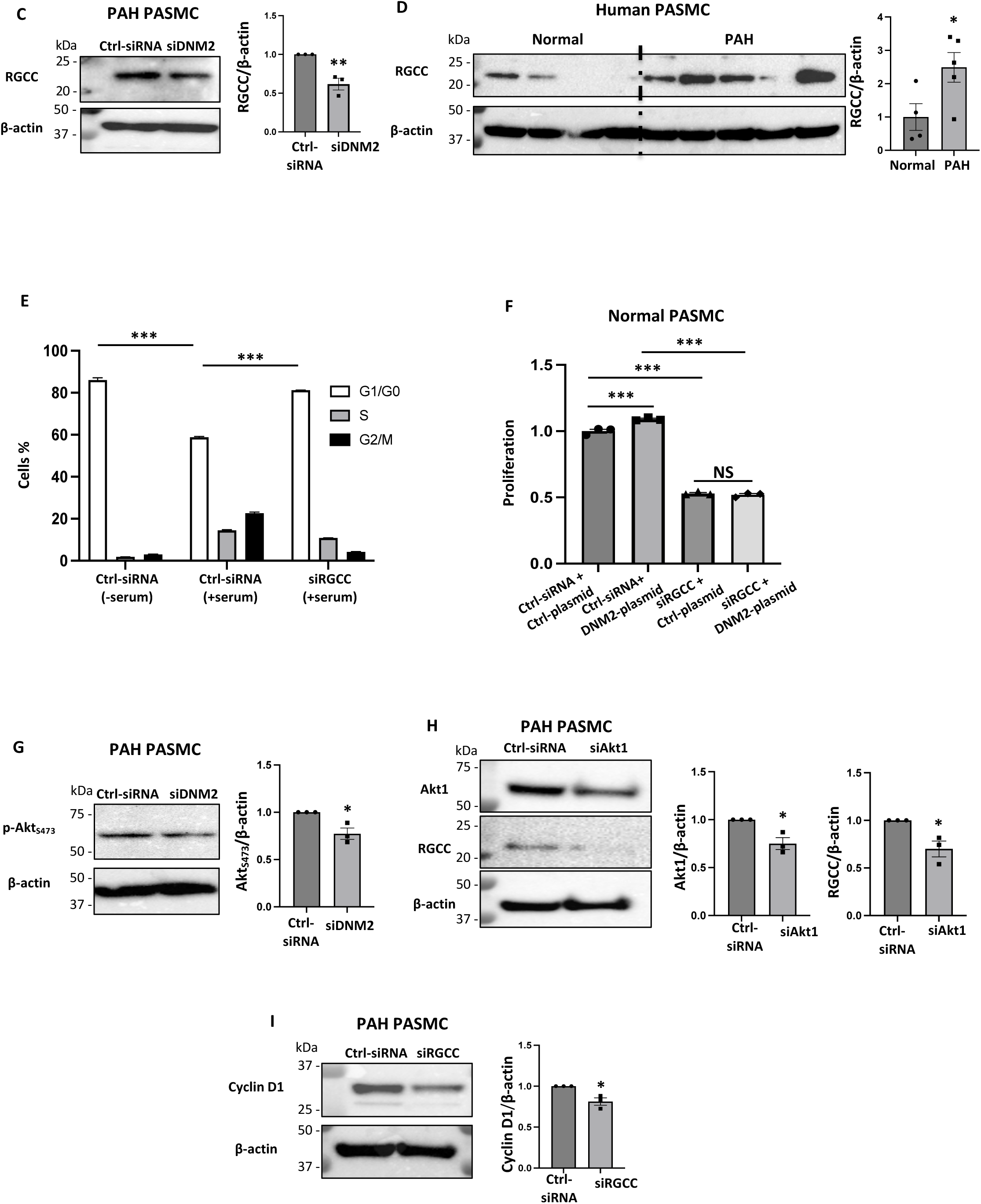
DNM2 regulates cell cycle via RGCC. A) Gene expression profiling in PAH PASMC following DNM2 silencing. A volcano plot showing the log_10_adjusted p-value, and the log_2_fold change of the dysregulated genes after knocking down DNM2 in 5 PAH PASMC cell lines. (green: upregulated genes and red: downregulated genes, blue: genes that were significantly regulated, but did not reach the 0.75-fold threshold). **B) The 28 enriched terms that include the phrase "Cell Cycle."** RGCC, highlighted in red, is shown to participate in 26 of the enriched terms given on the Y axis. These enriched terms were identified using a siDNM2 knockdown in human cells at a significance level of 0.1. **C) RGCC is downregulated in PAH PASMC following silencing of DNM2.** Representative image of immunoblot and densitometry of the expression of RGCC following silencing of DNM2. PAH PASMC were transfected with specified siRNA for 48 h. β-actin was used as the loading control (n=3 PAH cell lines) (\*\**P* < 0.01). **D) RGCC is upregulated in PAH PASMC.** Immunoblot and densitometry showing that RGCC is upregulated in PAH PASMC. PAH PASMC (n=5) vs control PASMC (n=4); β-actin was used as the loading control (\*\**P* < 0.01). **E) Silencing RGCC induces cell cycle arrest in the G1/G0 phase.** PAH PASMC was transfected with siRGCC for 24 h, serum starved for 24 h followed by serum stimulation for 24 h. Cell cycle analyses were performed by flow cytometry following propidium iodide (PI) staining (n=3 technical repeats) (\*\*\**P* < 0.001). **F) Augmenting DNM2 failed to induce proliferation in RGCC silenced PASMC.** Normal human PASMC were transfected with specified plasmids and siRNA. Cell proliferation was analyzed by EdU incorporation assay 72 h post-transfection (n=3 technical repeats) (\*\*\**P* < 0.001, NS: not significant). **G-I) DNM2 regulates the expression of RGCC via Akt**. **G) Silencing DNM2 inhibits Akt activation.** Representative image of immunoblot and densitometry showing the expression of p-Akt_S473._ PAH PASMC were transfected with specified siRNA. Cells were harvested for immunoblot analyses 48 h following transfection. β-actin was used as the loading control (n=3 PAH cell lines) (\**P* < 0.05). **H) Silencing Akt1 downregulates RGCC.** Representative image of immunoblots and densitometries showing the expression of Akt and RGCC. PAH PASMC were transfected with specified siRNA. Cells were harvested for immunoblot analyses 48 h following transfection. β-actin was used as the loading control (n=3 PAH cell lines) (\**P* < 0.05). **I) Silencing RGCC downregulates the expression of the cell cycle protein, Cyclin D1.** PAH PASMC were transfected with specified siRNA. Cells were harvested for immunoblot analyses 48 h following transfection. β-actin was used as the loading control (n=3 PAH cell lines) (\**P* < 0.05).

Silencing DNM2 reduced RGCC expression in PAH PASMC validating the RNA-seq result (Fig. 5C). Consistent with biologic relevance of RGCC in PAH, RGCC was upregulated in PAH PASMC (Fig. 5D) and, knocking down RGCC in PAH PASMC caused G1/G0 cell cycle blockade (Fig. 5E). Moreover, DNM2 augmentation failed to induce proliferation in normal PASMC in which RGCC had been silenced (Fig. 5F). Furthermore, it is reported that RGCC expression is regulated by Akt1^31^ and Akt1 is known to be activated in PAH PASMC^32^ (which we confirmed in our PAH PASMC, Supplementary Fig. 10). We show that silencing DNM2 inactivates Akt1, whereas silencing Akt1 downregulates RGCC expression. Thus, downregulation of RGCC following silencing of DNM2 is Akt-mediated (Fig. 5G-H). In addition, like siDNM2, siRGCC reduced cyclin D1 expression, indicating cell cycle blockade at the G1/G0 phase (Fig. 5I).

### Downregulation of miR-124-3p increases DNM2 in PAH via regulating STAT3 expression

We next addressed whether upregulation of DNM2 at the protein level is due to its reduced degradation or increased production. A cycloheximide chase study demonstrates that DNM2 protein is less stable in PAH PASMC than in normal PASMC, indicating the observed DNM2 upregulation is not due to impaired proteasomal degradation of DNM2 (Supplementary Fig. 11A). Next, we considered whether increased DNM2 in PAH results from decreased regulatory miRs and/or increased transcription. *In silico* analysis identified 12 miRs that putatively bind DNM2’s 3’-UTR (Supplementary Fig. 11B), of which, miR-124-3p^33^ and miR-204-5p were considered most promising because they are downregulated in PAH and can regulate cell proliferation and apoptosis^34, 35^. Most other miRs predicted to bind DNM2 are either upregulated or not reported to be dysregulated in PAH: e.g. miR-130-3p^36^, miR-301-3p^36^, miR-454-3p, miR-17-5p^37^, miR-20-5p^38^, and miR-211-5p^39^. In assessing the two candidate miRs, we found that augmenting miR-204-5p in PAH PASMC does not downregulate DNM2 expression, inhibit cell proliferation or induce apoptosis (Supplementary Fig. 11C-E). Thus, we focused on miR-124-3p^33^, and four other candidate miRs that are downregulated in PAH (miR-93-5p^40^, miR-106a-5p^41^, miR-133a-3p^42^ and miR-519-3p^43^). Because expression of miR-93-5p, miR-106a-5p and miR-133a-3p did not differ between normal and PAH PASMC they are excluded from further investigation (Supplementary Fig. 11F-H). miR-519-3p^43^ was downregulated in PAH PASMC but was excluded from further study because augmentation of miR-519-3p^43^ failed to downregulate DNM2 (Supplementary Fig. 11I-J). In contrast, miR-124-3p^33^ was decreased in human PAH PASMC (Fig. 6A) and augmenting miR-124-3p^33^ decreased DNM2 expression, inhibited mitochondrial fission and proliferation, and induced apoptosis in PAH PASMC (Fig. 6B-E).

**Figure 6.**
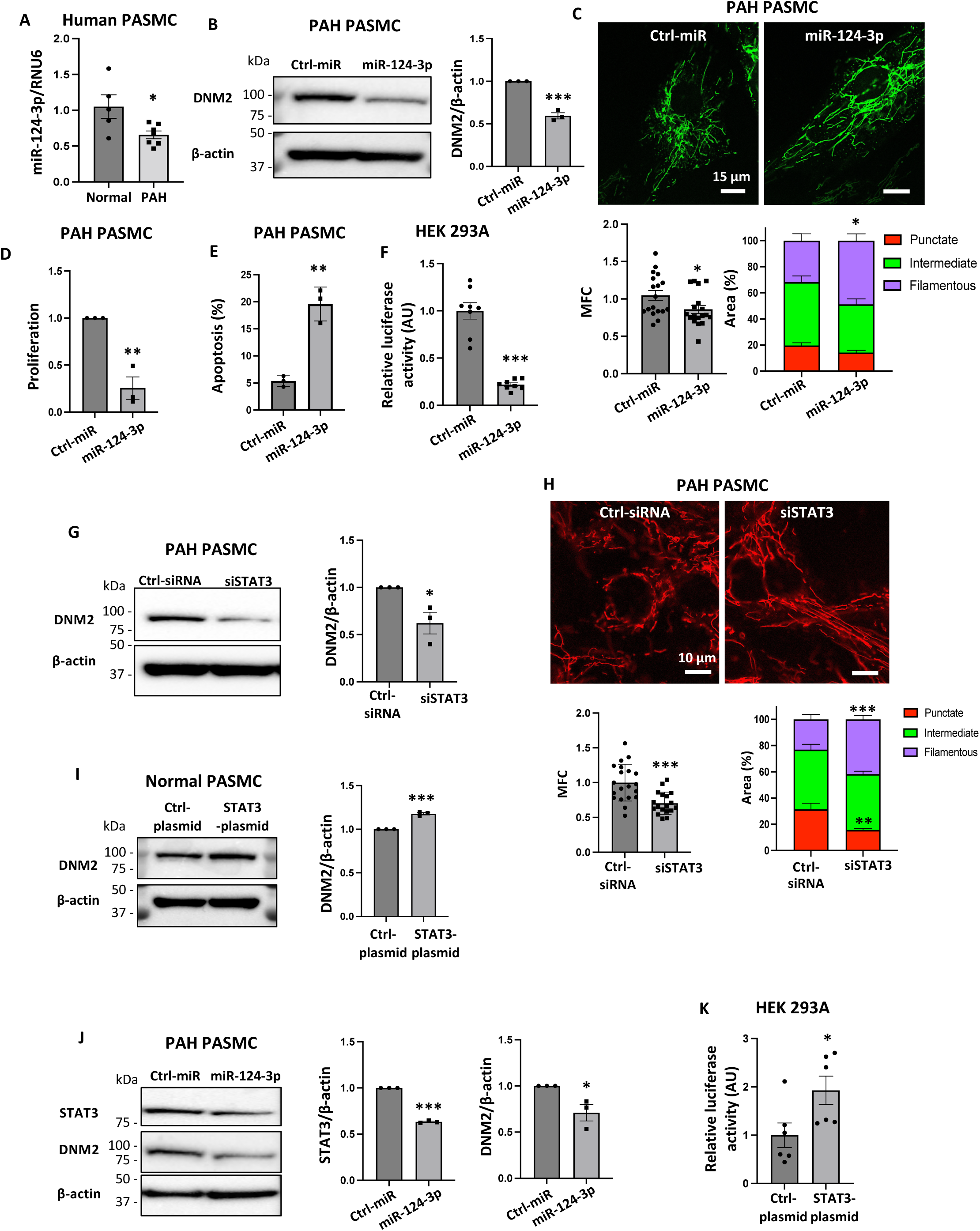
miR-124-3p and STAT3 are the regulators of DNM2 in human PAH PASMC. **A)** miR-124-3p is reduced in human PAH vs normal PASMC (n=5 PAH cell lines/group, qRT-PCR) (\**P* < 0.05). **B) Augmenting miR-124-3p downregulates the expression of DNM2 in PAH PASMC.** Representative immunoblot and densitometry showing the expression of DNM2 in PAH PASMC transfected with either ctrl-miR or miR-124-3p. Cell were harvested 48 h following transfection. β-actin was used as the loading control (n=3 PAH cell lines /group) (\*\*\**P* < 0.001). **C-E) Augmenting miR-124-3p inhibits mitochondrial fission and cell proliferation and induces apoptosis in human PAH PASMC**. **C)** Mitochondrial fragmentation was quantified by MFC and a machine learning algorithm was used to calculate the percentage area of punctate, intermediate and filamentous mitochondria of each image (n=18 cells/group) (\**P* < 0.05). **D)** PAH PASMC were transfected with either ctrl-miR or miR-124-3p. Cell proliferation was analyzed by EdU incorporation assay 72 h post-transfection (n=3 PAH cell lines/group) (\*\**P* < 0.01). **E)** PAH PASMC were transfected with either ctrl-miR or miR-124-3p. After 72 h the cells were labeled with Annexin V^FITC^ and PI and assessed by flow cytometry (n=3 technical repeats) (\*\**P* < 0.01). **F) miR-124-3p binds to 3’-UTR of DNM2.** miR-124-3p was found to repress the activity of luciferase reporter, indicating its binding to the 3’-UTR of DNM2 gene (n=8 technical repeats) (\*\*\**P* < 0.001). **G-H) Silencing STAT3 decreases DNM2 expression and inhibits mitochondrial fission in PAH PASMC.** PAH PASMC were transfected with either ctrl-siRNA or siSTAT3. **G)** Representative immunoblots and densitometry for the expression of DNM2 and **H)** Confocal imaging after loading the cells with TMRM were performed 48 h post-transfection with specified siRNA. β-actin was used as the loading control for immunoblot analyses. Mitochondrial fragmentation was quantified using MFC, and a machine learning algorithm was used to calculate the percentage area of punctate, intermediate and filamentous mitochondria of each image (n=3 PAH cell lines/group for DNM2 expression and n=18-20 cells/group for mitochondrial morphology) (\**P* < 0.05, \*\**P* < 0.01, \*\*\**P* <0.001). **I) Overexpressing STAT3 increases DNM2 expression in normal PASMC.** Representative immunoblot and densitometry showing the expression of DNM2. Normal PASMC were transfected with specified plasmids. Cells were harvested for immunoblot analyses 48 h following transfection. β-actin was used as the loading control (n=3 normal PASMC cell lines) (\*\*\**P*<0.001). **J) DNM2 and STAT3 expressions are regulated by miR-124-3p.** Representative immunoblots and densitometries for the expressions of STAT3 and DNM2. PAH PASMC were transfected with specified siRNA and the cells were harvested for immunoblot analyses 48 h following transfection (n=3 PAH cell lines/group) (\**P* < 0.05, \*\*\**P* < 0.001). **K) STAT3 binds to the promoter region of DNM2.** STAT3 was found to increase the activity of luciferase reporter, indicating its binding to the promoter region of DNM2 gene (n=6 technical repeats) (\**P* < 0.05).

The interaction between miR-124-3p and the 3’-UTR of DNM2 mRNA was confirmed by a luciferase reporter assay in which luciferase reporter, fused to the 3’-UTR of the DNM2 gene, interacted with a co-transfected miR-124-3p mimic reducing luciferase activity (Fig. 6F). These data strongly support the regulation of DNM2 by miR-124-3p.

However, miR-124-3p also negatively regulates signal transducer and activator of transcription 3 (STAT3)^44^, and STAT3 can positively regulate DNM2 expression^45^. Therefore, we also assessed whether increased expression of STAT3 in PAH also induces upregulation of DNM2. siSTAT3 downregulated DNM2 and inhibited mitochondrial fission in PAH PASMC (Fig. 6G-H) whilst augmenting STAT3 in normal PASMC upregulated DNM2 (Fig. 6I). Thus, STAT3 is a positive regulator of DNM2. Next, we investigated whether pathologically reduced miR-124a-3p levels directly increase DNM2 expression or do so via augmenting STAT3 (or both). Our results show that augmenting miR-124-3p downregulated both STAT3 and DNM2 expression in PAH PASMC (Fig. 6J). Furthermore, co-transfection of HEK293A cells with STAT3-plasmid and the luciferase reporter construct fused to the promoter region of *DNM2* resulted in increased luciferase activity, confirming the binding of the STAT3 to *DNM2’s* promoter region (Fig. 6K). These results indicate that the expression of DNM2 in PASMC is regulated by both miR-124-3p and STAT3 and show reduced miR124-3p can upregulate STAT3.

### Silencing DNM2 regresses PAH in MCT-PAH rats

Rats were allowed to develop PAH prior to administration of siDMN2 or control siRNA by nebulization (Fig. 7A and Supplementary Fig. 12). The knock-down efficiency of siDNM2 in small PAs was confirmed by immunofluorescence (Fig. 7B). siDNM2 treatment regressed vascular obstruction, as quantified by a decrease in medial thickness in MCT-PAH rats (Fig. 7C). Furthermore, siDNM2 treatment improved hemodynamics as evidenced on RHC by decreased right ventricular systolic pressure (RVSP) (Fig. 7D), a significant increase in cardiac index (CI) and fall in pulmonary vascular resistance index (PVRI) (Fig. 7E-F). Moreover, siDNM2 caused a marginally significant improvement of Fulton index (Fig. 7G). siDNM2 treatment did not result in any hematological, hepatic or renal toxicity in MCT-PAH rats (Supplementary Fig. 13A-K).

**Figure 7:**
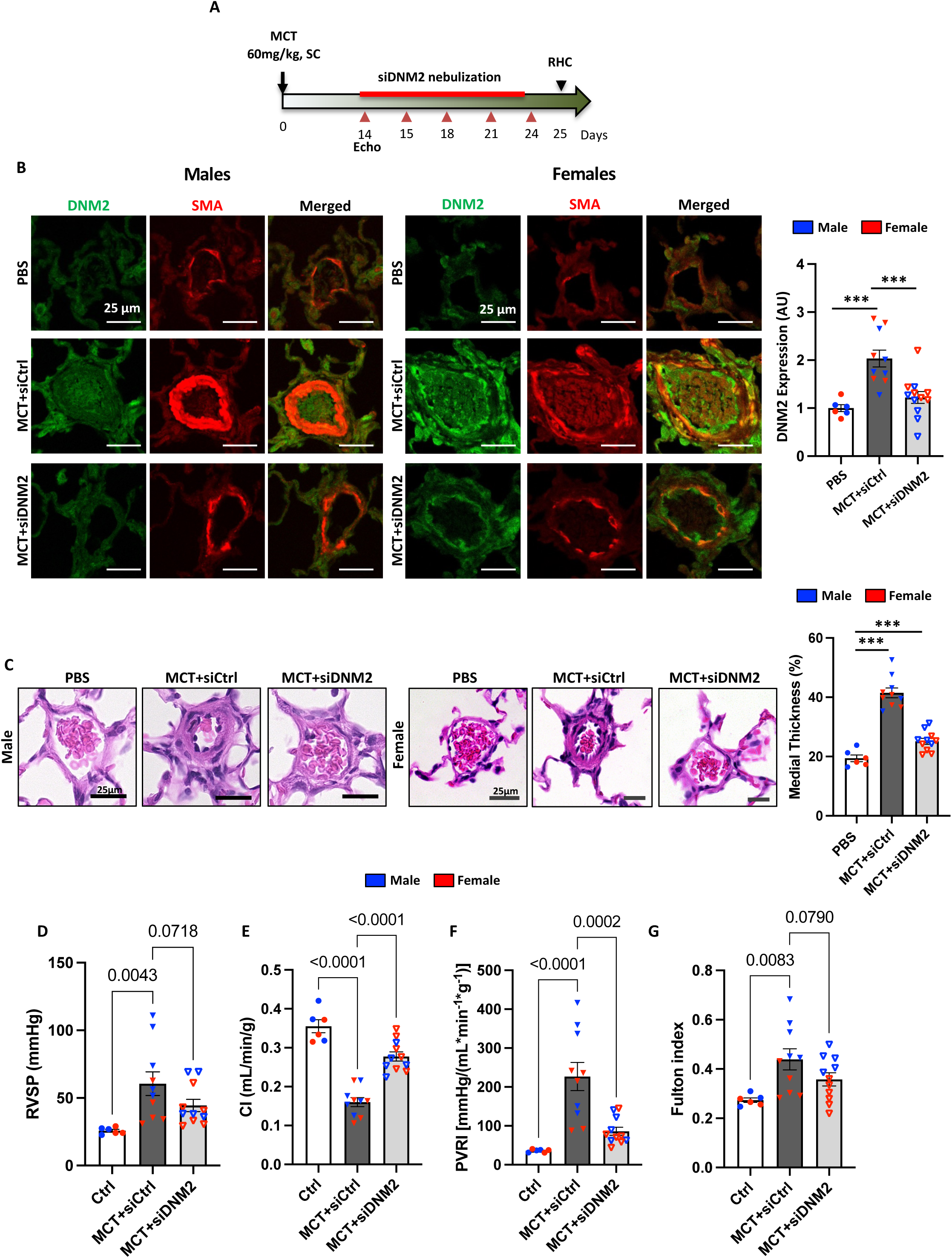
Therapeutic relevance of the silencing DNM2 via siRNA nebulization in the monocrotaline-PAH preclinical model. **A)** Study design of the animal study. **B)** Representative cross-sectional images of small pulmonary arteries of both male (n=3-6 animals/group) and female (n=3-6 animals/group) MCT-PAH rats showing decreased DNM2 expression in the media of small PAs with siDNM2 treatment (\*\*\**P* < 0.001). **C)** Representative images of pulmonary vascular histology showing siDNM2 reduced medial thickness of small PAs in both males (n=3-5 animals/group) and females (n=3-5 animals/group) MCT-PAH rats. H&E staining. Scale bar: 25 μm. (\*\*\**P* < 0.001). **D-F)** siDNM2 treatment improved pulmonary arterial hypertension and RV function, which is impaired in MCT-PAH rats. Compared to PBS rats, MCT-PAH rats had **D)** increased right ventricular systolic pressure (RVSP), **E)** decreased cardiac index (CI), and **F)** increased pulmonary vascular resistance index (PVRI), as determine by closed-chest right heart catheterization. siDNM2 treatment was effective in **D)** decreasing RVSP, **E)** improving CI while **F)** decreasing PVRI. **G)** The RV hypertrophy induced by MCT in MCT-PAH rats, indicated by a significantly increased Fulton index, is also reversed by siDNM2 treatment (n=6-11 animals/group).

## Discussion

This study offers six important findings: 1) DNM2-mediated fission depends on Drp1. Indeed, DNM2 translocates to mitochondria when its GTPase domain binds to Drp1. 2) Pathological upregulation of DNM2 occurs in human PAH and rodent PAH models. 3) Downregulating DNM2 in PAH PASMC inhibits mitochondrial fission, blocks cell cycle progression and triggers apoptosis. 4) DNM2 exerts positive regulatory effects on RGCC, a cytosolic activator of cyclin dependent kinases, which is a molecular link between mitotic fission and cell cycle progression. We show that DNM2 regulates the expression of RGCC by positively regulating Akt1. 5) miR-124-3p deficiency and STAT3 activation together upregulate DNM2 in human PAH. 6) Nebulized siDNM2 regresses established MCT-PAH. Thus, DNM2 is not only relevant to fundamental mechanisms of mitochondrial fission but also links fission and cell cycle regulation. On a translational level these findings have relevance to PAH, as DNM2 upregulation contributes to the pathogenesis of this hyperproliferative vascular disease.

Prior studies of DNM2’s role in mitochondrial dynamics are limited in number and their findings are non-concordant, with one report supporting^46^ and another refuting^47^ a role of DNM2 in mitochondrial fission. Neither of these earlier studies examined DNM2’s role in human disease. Lee et al concluded that Drp1 lacks the dynamic constriction range to complete mitochondrial fission^46^. They proposed that the final division event is carried out by DNM2, which can constrict lipid membranes to <10nm^46, 48^. They showed that DNM2 is recruited to ER-induced mitochondrial fission sites and, in HeLa cells, found that siDNM2 causes mitochondrial hyperfusion, similar to cells lacking Drp1^46^. Contradicting Lee’s findings, Fonseca et al found that triple knockdowns of all dynamin isoforms (1-3) did not alter mitochondrial fission^47^. Our study supports and extends the work of Lee et al.

Currently, the established function of DNM2 is the scission of plasma membrane vesicles during endocytosis^49^. DNM2 also forms vesicles from the Golgi apparatus and regulates vesicle trafficking and the dynamics of microtubules and the actin cytoskeleton^50–52^. Furthermore, inhibiting DNM2 in cancer reduces cell proliferation and inducing apoptosis in cervical cancer^53^, prostate cancer^54^, non-small-cell lung cancer^55^, glioblastoma^56^, and chronic myeloid leukemia^57^. While inhibitors of mitochondrial fission (or enhancers of fusion) regress human cancers of various types^17, 18^, prior studies of DNM2 in cancer did not investigate DNM2 as a mediator of mitochondrial fission nor implicate mitochondrial biology in the preclinical efficacy of DNM2 inhibition.

We demonstrated that the critical role of DNM2 in executing mitochondrial fission (Fig. 2A-E), is achieved through a partnership with Drp1 (Fig. 3A-E). This conclusion is supported by our observation of increased length of the constriction sites following DNM2 silencing, indicating mitochondrial fission is impaired by the loss of DNM2 (Fig. 2G). Furthermore, in comparison to normal PASMC, PAH PASMC exhibits tighter constrictions at fission sites, as evidenced by a decrease in the length and residual thickness of the mitochondrial at constriction sites. We interpret this as evidence that greater mitochondrial DNM2 expression promotes the excess fission seen in PAH PASMC (Fig. 2F). Our observation of Drp1-DNM2 interaction (by immunoprecipitation) and closer proximity of these molecules at fission sites in PAH versus normalPASMC (by super-resolution STED microscopy) is consistent with the findings of Lee et al, who reported incomplete Drp1-marked fission (stalled super-constrictions) at mitochondrial fission sites in DNM2-depleted cells^46^. There is no significant difference in maximal mitochondrial diameter between normal and PAH PASMC (each ∼400nm on confocal imaging, Fig. 2F); however, the true diameter of mitochondria (measured using STED, which has 5-fold higher resolution) is ∼200nm (for both normal and PAH mitochondria), attesting to the importance of super resolution imaging in assessment of mitochondrial dynamics (Supplementary Fig. 5E). Therefore, the observed reduced length and thickness of the mitochondrial constriction sites in PAH PASMC is not due to any pre-existing differences in baseline mitochondrial thickness, rather it reflects the enhanced narrowing caused when DNM2 interacts with Drp1 on the mitochondria (Fig. 3J-K).

Like Drp1, DNM2 lacks mitochondrial-targeting domains. Interestingly, Drp1 conveys DNM2 to the mitochondria. Our data demonstrate that DNM2 interacts with Drp1 in the cytosol as well as at the site of mitochondrial fission (Fig. 3A-D) and that silencing Drp1 inhibits translocation of DNM2 to the mitochondria (Fig. 3E). Furthermore, augmenting DNM2 in Drp1 conditional knockout MEFs, increased fission when Drp1 was present; however, when Drp1 was conditionally knocked out, DNM2 no longer caused fission (Fig. 3F-G). Thus, DNM2 mediated mitochondrial fission is Drp1-dependent. Furthermore, a structure-function study revealed that a DNM2 construct lacking GTPase domain (ΔGTPase) failed to localize to mitochondria or interact with Drp1, and thus failed to induce mitochondria fission. These findings highlight the critical role of DNM2’s GTPase domain in binding Drp1 and targeting mitochondria (Fig. 3H-J). Conversely, the constructs carrying the GTPase domain of DNM2 were sufficient to co-localize and interact with Drp1 and translocate to mitochondria (Fig. 3J-K and Supplementary Fig. 6B), further highlighting the crucial role of this domain in DNM2’s mitochondrial targeting and fission. These findings emphasize an orchestrated action of the Drp1-DNM2 complex in executing mitochondrial fission.

We observed that the basal expression of DNM2 on normal mitochondria is low, and normal PASMC can be rendered hyperproliferative by heterologous overexpression of DNM2. However, in PAH, there is pathological upregulation of DNM2 (Fig.1A-F), which increases mitotic mitochondrial fission, leading to hyperproliferation and apoptosis resistance of PAH PASMC (Fig. 2A-E and Fig. 4A-H). Interestingly, among the isoforms of Dynamin family, only DNM2 is upregulated in PAH, highlighting its critical role in disease pathogenesis (Fig. 1A-F and Supplementary Fig. 2A-B). This preferential importance of the DNM2 isoform in PAH is consistent with the elevated expression of DNM2 in many cancers^58^.

The importance of DNM2 in the pathogenesis of PAH was further supported by our observation that silencing DNM2 restored mitochondrial network fusion, inhibited proliferation by causing a cell cycle blockade at G1/G0 phase, and induced apoptosis in PAH PASMC (Fig. 2A-E and Fig. 4A-G). Conversely, augmenting DNM2 mimicked a PAH phenotype in normal PASMC by inducing mitochondrial fission and cell proliferation (Fig. 2C and Fig. 4H).

Further supporting that DNM2 inhibition causes cell cycle arrest at the G1/G0 phase, siDNM2 decreases the expression of the cell cycle proteins CDK4 and cyclin D1 (Fig. 4C-D) and increases in the expression of the CDK4 inhibitor p27^kip1^ (Fig. 4E). We previously reported that inhibition of CDK4 and upregulation of p27^kip1^ occurs when Drp1 receptor proteins, MiD49 and MiD51, were silenced in human PAH PASMC^23^. This suggests that inhibiting mitochondrial fission, whether through inhibition of Drp1, Drp1 binding partners, or DNM2, induces changes in cell cycle mediators (downregulation of positive regulators and upregulation of inhibitors) which mediate cell cycle blockade^23^.

In addition, we identified a new mechanism(s) that connects DNM2 to cell cycle progression. After silencing DNM2 in PAH PASMC, using RNA-seq analysis we found that RGCC is the most downregulated gene (Fig. 5A and Table 2). RGCC (also known as RGC-32) is a cytosolic protein that activates cyclin dependent kinases potentially playing a key role in regulating the cell cycle. RGCC is known to be involved in smooth muscle proliferation and migration, angiogenesis, and cancer development^31, 59^. Like DNM2, RGCC expression is upregulated in PAH PASMC (Fig. 5D). and silencing RGCC expression also causes cell cycle arrest in PAH PASMC at the G1/G0 phase (Fig. 5E). Moreover, in RGCC-silenced PASMC, DNM2 failed to induce proliferation, indicating that DNM2 regulates cell proliferation through RGCC (Fig. 5F). This is the first demonstration that RGCC is a key downstream mediator in DNM2-mediated regulation of cell cycle progression. Furthermore, Akt1 which is known to regulate RGCC expression^31^, is activated in PAH PASMC (Supplementary Fig. 10). We also showed that silencing DNM2 reduces Akt1 activation (Fig. 5G). Given that the expression of RGCC is regulated by Akt1 (Fig. 5H), and that Akt1 itself is regulated by DNM2, we conclude that DNM2 regulates the expression of RGCC by regulating Akt1.

The increase in DNM2 expression in PAH PASMC is due both to reduced miR-124-3p and increased expression of the transcription factor, STAT3. Indeed, miR-124-3p deficiency also elevates STAT3 expression, highlighting redundancy in this pathological pathway (Fig. 8). In PAH PASMC restoring miR-124-3p downregulates DNM2 expression, inhibiting mitochondrial fission, suppressing proliferation and inducing apoptosis (Fig. 6C-E). Further molecular evidence that miR-124-3p regulates DNM2 is the demonstration of direct binding of miR-124-3p to the 3’-UTR of DNM2 (Fig. 6F). Dysregulation of miR-124-3p has also been implicated in causing metabolic and proliferative abnormalities in pulmonary artery endothelial cells of PAH patients via PTBP1 and the PKM1/PKM2 glycolytic, Warburg metabolic pathway^8^. How the miR-124/PTBP1/PKM axis and miR-124/DNM2 pathways interact merits future study.

**Figure 8:**
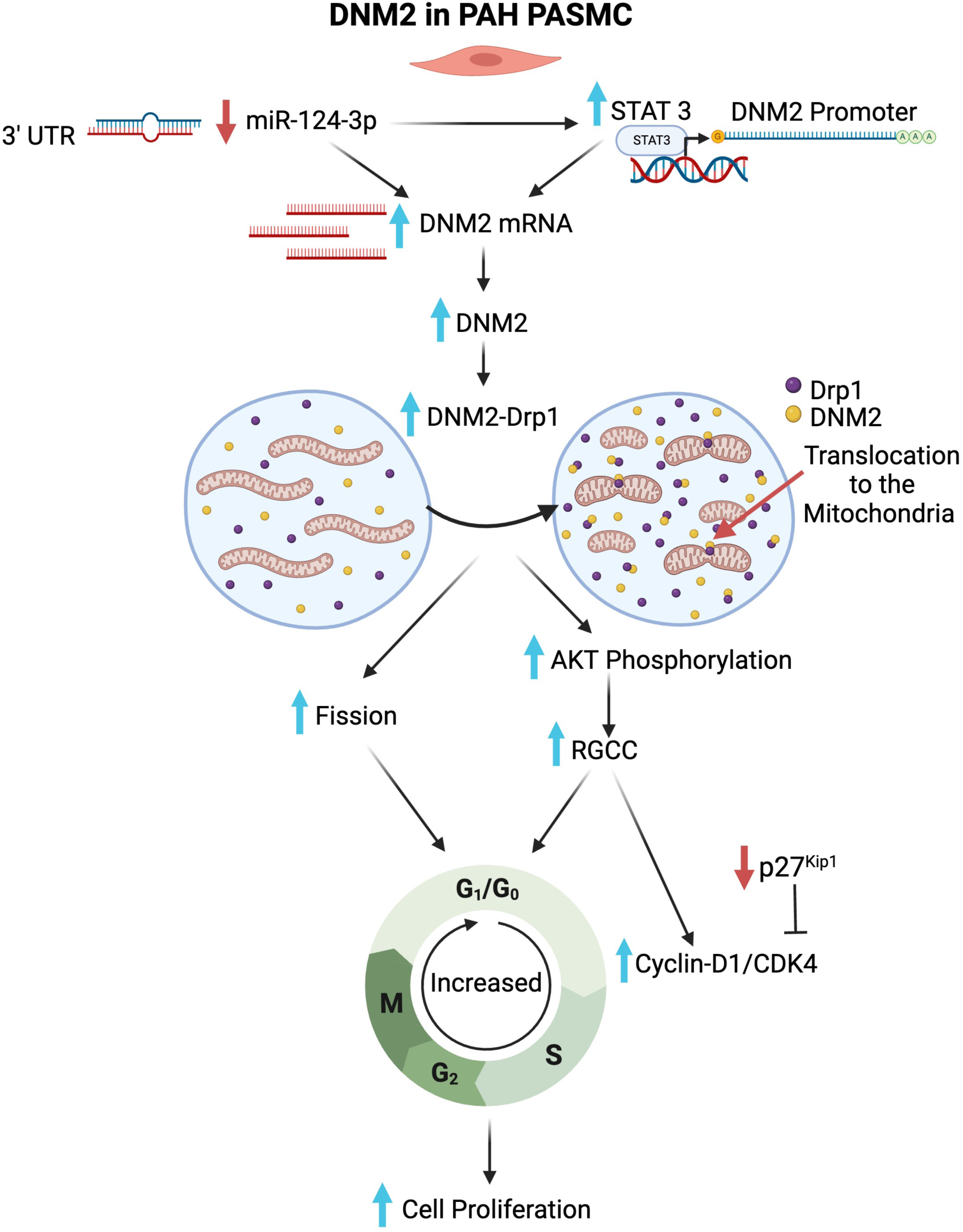
Representation of the proposed miR-DNM2 pathway in PAH PASMC. Decreased expression of miR-124-3p and increased expression of STAT3 in PAH patients leads to increased expression of the DNM2 which in turn increases mitotic fission and promotes cell proliferation via a mechanism that is dependent on activation of Akt and RGCC.

DNM2 expression is also positively regulated by STAT3^45^, a regulator of cell proliferation and apoptosis, which is constitutively activated in PAH^60–62^. Evidence implicating STAT3’s contribution to upregulating DNM2 expression is the demonstration that silencing STAT3 in PAH PASMC downregulated DNM2 expression and inhibited mitochondrial fission; whilst augmenting STAT3 in normal PASMC increased DNM2 expression (Fig. 6G-I). A strength of our conclusion of the role of STAT3 in regulating DNM2 is the demonstration of a direct interaction of STAT3 with DNM2’s promoter region (Fig. 6K). These findings suggest that STAT3 can directly control DNM2 transcription in PAH PASMC; however, miR-124-3p can negatively regulate STAT3 expression^44^. Thus pathologically low miR-124-3p levels can directly increase DNM2 expression (Fig. 6F) or do so indirectly by increasing the expression of STAT3 (Fig. 6J). We conclude that the elevated DNM2 expression in PAH PASMC is a combined consequence of an epigenetic mechanism (decreased miR-124-3p expression) and a transcriptional mechanism (increased STAT3).

We previously demonstrated that creating a persistent state of mitochondrial fusion, whether by inhibiting Drp1, augmenting mitofusin 2, or targeting Drp1 adaptors (MiD49/51), suppresses cell proliferation and regresses PAH and cancers^17, 18, 23, 63^. Consistent with this, we now show that inhibiting DNM2 causes sustained mitochondrial fusion in PAH PASMC resulting in the suppression of cell proliferation and induction of basal apoptosis. These findings offer a strong rationale for therapeutically targeting DNM2 in PAH in vivo, Indeed, nebulizing siDNM2 in an MCT-induced PAH rat model regressed PAH, in both sexes, and improved some dysregulated hematological, hepatic, and renal parameters (Fig. 7B-G, Supplementary Fig. 13A-K).

### Limitations

This study has limitations. First, we tested the therapeutic efficacy of silencing DNM2 only in MCT-PAH model. However, DNM2 upregulation was noted in the Su/Hx PAH models, as well as in patients with PAH. Second, the diagnostic and prognostic values of DNM2 and miR-124-3p in PAH were not evaluated in a patient cohort, warranting future investigation.

In conclusion, DNM2 participates in the terminal step of mitochondrial fission (Fig. 8). Epigenetic and transcriptional upregulation of DNM2 increase mitotic fission in PAH PASMC and promotes pulmonary vasculopathy. The underlying mechanism that connects DNM2 and fission to cell cycle progression involves RGCC. The miR-124-3p-STAT3-DNM2-Drp1-RGCC pathway offers potential therapeutic targets in PAH.

## Data Availability

All data produced in the present study are available upon reasonable request to the authors.

## Author contributions

Dasgupta A., Chen KH. and S. L. Archer designed the research protocol and proposed the hypothesis; A. Dasgupta, KH. Chen, D. Wu, Yerramilli VS, P.D.A. Lima, A. Martin, J.D. Mewburn, L. Tian, R. Al-Qazazi, O. Jones, P. Colpman, L. Jefferson, C. Noordhof, I. Emon C.CT. Hindmarch, performed the research; A. Dasgupta, KH. Chen, D. Wu, P. D.A. Lima B.P. Ott, and C.CT. Hindmarch analyzed the data. A. Dasgupta, and S. L. Archer wrote the manuscript; S. L. Archer supervised the experiments. S. L. Archer and A. Dasgupta acquired the funding.

## Acknowledgements

We acknowledge Dr. Sébastien Bonnet Laval University, Quebec City, Quebec, Canada for providing cells and human lung sections used in this study. We also acknowledge the Translational Institute of Medicine (TIME) and Queen’s Cardiopulmonary Unit (QCPU) at Queen’s University for their support.

