## Supplementary Method for "Role of Dynamin 2 in mitochondrial fission and cell cycle regulation: *Dysregulation of a miR-124-3p-STAT3-DNM2-Drp1-RGCC pathway connects fission and cell proliferation in pulmonary arterial hypertension*"

**siRNA and primers:**

The siRNA duplex specific for human DNM2 (#SR301243), RGCC (#SR309156), siDNM3 (#SR308690), Mfn2 (#SR322626) and STAT3 (#SR321907) were purchased from OriGene (Rockville, MD, USA). siAkt1 (#AM16078) was purchased from Ambion (Austin, Tx, USA). Control siRNA (#51-01-14-04), siMFF (HSC. RNAI. N020194.12), Fis1 (HSC. RNAI. N016068.12.5) siMiD49 (HSC. RNAI.N001144900.12.2) and siDNM1 (HS.R1.DNM1.13.2) were obtained from Integrated DNA Technologies (Coralville, IA, USA).

**RNA sequencing**

Libraries for RNAseq analysis were constructed using the Lexogen 3' kit according to manufacturers protocols (Lexogen, Austria), and validated using Qubit 4 (Thermo). Successful libraries were pooled and diluted to an equimolar concentration resulting in a total pool of 4nM. Dilution and denaturing were performed according to Illumina protocols to result in a 2.2pM (optimized for the Lexogen 3' kit). Single-end sequencing of 75 cycles was performed on an Illumina Nextseq550 using a High Output v2 reagent cartridge with each sample having a minimum depth of 5 million reads. Sequence files were demultiplexed, and processed using fastqc [1] and multiqc prior to trimming with BBDuk [2], alignment to the human GRCh38 genome with Star

[3], and counting with HT-Seq Count [4]. Differential analysis was performed using a pair-wise approach and implemented in DESeq2[5] package (v1.44) in R (v4.4.0)[6] and they were included in the GO:BP (Gene Ontology Biological Process) analysis using gprofiler2 (v.0.2.3).

**Plasmid construction:**

For cloning DNM2 constructs (See Supplemental Material) pCMV6-Entry (#PS100001) and pCMV6-AC-GFP (#PS100010) mammalian expression vectors were purchased from OriGene (Rockville, MD, USA) and were used as the backbones. The sequence of primers used for cloning are: DNM2 GTPase domain (sense): 5'-CCCCCGCGATCGCC ATGGGCAACCGCGGGATGGAAG-3', (antisense) 5'-CCCCCACGCGTGCC TCTTCTCAACGGGAGCAAC-3'. DNM2-ΔGTPase domain (sense): 5'-CCCCCGCGATCGCCATGGGCTACATTGGCGTGGTGAACC-3', (antisense): 5'-CCCCCACGCGT GTCGAGCAGGGATGGCTCGGC-3'. DNM2 full length (sense): 5'-CCCCCGCGATCGCCATGGGCAACCGCGGGATGGAAG-3', (antisense) 5'-CCCCCACGCGT GTCGAGCAGGGATGGCTCGGC-3'.

**DRP1 KO MEFs:**

Drp1 KO MEFs was a kind gift from Dr. Gerald Dorn II (Washington University, St Louis, USA) and were generated from Drp1$^{loxp/loxp}$ embryos[7]. The MEFs were maintained in DMEM supplemented with 10% FBS, 100 U/ml penicillin, 100 µg/ml streptomycin, 2 mM glutamine and 1x non-essential amino acids.

**Immunofluorescence staining:**

Briefly, rat lung sections were labelled with antibodies specific for smooth muscle actin (SMA) (#1497-60-82, eBioscience, San Diego, CA, USA), to identify the media, and DNM2 (#ab3457, Abcam Cambridge, MA, USA). Immunofluorescence staining was performed on normal and PAH PASMCs on glass coverslips after fixing with 4% Paraformaldehyde and permeabilization with 0.5% Triton X in PBS. The cells were stained with antibodies specific for DNM2 and Drp1(# 61113 BD Biosciences) followed by corresponding secondary antibody after which they were mounted on glass slides with Prolong Diamond antifade mountant with DAPI. The cells are loaded with MitoTracker Deep Red (excitation 644 nm and emission 665 nm (Thermofisher Scientific, Waltham, CA, USA) to stain Mitochondria prior to being fixed.

**Histology Methods**

The tissue was immersed in 10% neutral buffered formalin for 24 hours before processing, on a Leica HistoCore Pearl tissue processor, embedded in paraffin wax. Tissue sections were then cut on a Leica MULTICUT microtome at 4 microns thickness, and stained with hematoxylin and eosin.

**Western blotting:**

The whole-cell lysates were prepared by lysing the cell pellets with lysis buffer (Cell Signaling Technologies, Beverly MA, USA). 40-60 µg of cell lysates were analyzed for immunoblot analyses on 4–12% NuPAGE gels (Life Technologies, Carlsbad, CA, USA). Polyvinylidene difluoride (PVDF) membrane (Life Technologies, Carlsbad, CA, US) was used to electrotransfer the proteins and specific proteins were detected by using indicated antibodies and by the ECL-Plus Western Blotting Detection System (GE Healthcare, Piscataway, NJ, USA).

**Antibodies:**

Antibodies against DNM2 (ab3457) and Mfn2 (ab56889) were purchased from Abcam (Cambridge, MA, USA). Antibodies specific for DNM1(PA1-660) and DNM3 (PA1-662) were purchased from Invitrogen (Waltham, MA USA). Antibody specific for Drp1 (611738) was obtained from BD Transduction Laboratories (Becton Drive, NJ, US). Antibodies against MiD49 (16413-1-AP), Fis1 (10956-1-AP), MFF (17090-1-AP) and CDK4 (11026-1-AP) were purchased from Proteintech (Tucson, AZ, USA). Antibodies against pan Akt (4691), p-Akt (ser473) (9271), Cyclin D1 (2922), $p27^{Kip1}$ (3686), Bak (6947), were obtained from Cell Signaling Technology (Beverly, MA, USA). Antibody against RGCC (NBP2-93098) was purchased from Novus Biologicals (Centennial, CO, USA).

**Immunoprecipitation:**

Cell signaling lysis buffer (Cell Signaling Technologies, Beverly, MA, USA) supplemented with a protease and phosphatase inhibitor cocktail (Thermo Fisher Scientific, Waltham, MA, USA) was used to lyse the cell pellets. The cell lysates were incubated with anti-Drp1 antibody at 4$^{o}$C overnight to perform immunoprecipitation reactions. The A/G-PLUS agarose beads (Santa Cruz Biotechnology, Dallas, TX, USA) were used to recover the immune complexes. The immune complexes were washed with buffer and the beads were boiled in NuPAGE sample buffer (Invitrogen, Carlsbad, CA, USA) to elute the bound proteins and the immunoprecipitated proteins were analyzed by immunoblotting [8].

To perform immunoprecipitation reactions with recombinant proteins, recombinant Drp1 (TP321708) and DNM2 (TP323585) proteins were obtained from OriGene (Rockville, MD, USA). Immunoprecipitation reactions were performed with anti-Drp1 antibody following the protocol stated above.

**Cell transfection:**

For siRNA treatment, PAH PASMC were grown to 60-80% confluence and then transfected with 25 picomole of siRNA using the Lipofectamine® *RNAiMAX* Transfection Reagent (Life Technologies, Carlsbad, CA). The knockdown efficiency was assessed after 48 hours using qRT-PCR (Bio-Rad, Hercules, CA, USA) and immunoblotting. For plasmid overexpression, the cells were transfected with 2.5 μg specified plasmid using Lipofectamine 3000 (Life Technologies, Carlsbad, CA).

**Cell cycle analysis:**

The PAH PASMC were transfected with specified siRNA. The cells were then synchronized at G1/G0 phase for 24 hours by serum starvation and then stimulated with 15% FBS medium for 24 hours to release them from synchronization. Following 24 hours of stimulation, the cells were harvested and suspended in 300 µl PBS, and fixed with 70% ethanol at -20°C. The cells were then washed twice with ice cold PBS and incubated with 500 µl PI/RNase staining buffer (BD Biosciences, Franklin Lakes, NJ, USA) at room temperature for 15 minutes. The samples were analyzed for DNA content by detecting the PI binding to DNA using a fluorescence-activated cell sorter (Sony SH-800, San Jose, CA, USA).

**Apoptosis Assay:**

The PAH PASMC were grown to 80-90% confluence in 15% FBS medium and transfected with 25 picomole of ctrl-siRNA, siDNM2, ctrl-miR or miR-124-3p using Lipofectamine® RNAiMAX. Transfection Reagent (Life Technologies, Carlsbad, CA). 72 hours following transfection, the cells were stained with the Alexa Fluor 488 Annexin V/Dead Cell Apoptosis Kit (Life Technologies, Carlsbad, CA, USA) following the manufacturer's instruction and analyzed for apoptosis using Flow cytometry (Sony SH800S, Sony Biotechnology, San Jose, CA, USA).

**Confocal and Stimulated Emission Depletion (STED) Imaging of Live Cells:**

Live cells were cultured in glass-bottom dishes (MatTek Corporation, Ashland, MA, USA) and transfected with specified siRNAs or miRs if applicable. 48 h following transfection, the cells are loaded with tetramethylrhodamine, a mitochondrial potentiometric dye (TMRM; 20 nM, Molecular Probes, Eugene, OR, USA, excitation 561 nm, emission > 575 nm) or with a membrane potential independent mitochondrial probe, MitoTracker Green (excitation 491 nm and emission 560 nm, Thermofisher Scientific). The cells were imaged with a Leica SP8 laser scanning confocal microscope using a 1.40NA, 63x oil immersion objective (Leica, Planapo, Wetzlar, Germany). STED imaging of cells loaded with MitoTracker Green was performed using a 1.40 NA 100x oil immersion STED objective (Leica) supplemented with a 595 nm depletion laser.

**Confocal and STED Imaging of Fixed Cells:**

The cells fixed on glass coverslips were imaged with a Leica SP8 laser scanning confocal microscope using a 1.40NA, 100x oil immersion objective (Leica, Planapo, Wetzlar, Germany). STED imaging was performed using 595 nm and 660 nm depletion lasers.

**Quantification of mitochondrial morphology and networking:**

Mitochondrial fission was quantified using two previously validated metrics that measure mitochondrial structure: mitochondrial fragmentation count (MFC) and a machine learning algorithm, as described[9, 10].

A reduction of MFC indicates a more fused mitochondrial network. Mitochondrial networking was further quantified using a metric called the mitochondrial networking factor (MNF), which quantifies the diffusion of mitochondrial targeted, photoactivatable green fluorescent protein (GFP) along the mitochondrial matrix, as previously described[9, 10]. A higher degree of mitochondrial network fusion was indicated by a higher MNF.

**miR-124-3p and STAT3 binding luciferase reporter assay:**

The binding of miR-124a-3p to the 3'-UTR of DNM2's mRNA was validated using a luciferase binding assay as previously described[9, 11]. Briefly, the HEK293A cells were co-transfected with a reporter plasmid containing the 3'-UTR of DNM2 (OriGene, Rockville, MD, USA) together with miR-ctrl or miR-124-3p using Lipofectamine 3000 (for plasmid transfection) and RNAiMAX (for miR transfection) transfection reagents (Life Technologies, Carlsbad, CA). Luciferase activity assay was performed as previously described[9, 12].

The binding of STAT3 to DNM2's promoter region was validated using a promoter binding assay. Briefly, the HEK293A cells were co-transfected with a reporter plasmid containing the promoter region of DNM2 (OriGene) together with either a control or a STAT3 plasmid (OriGene) using

Lipofectamine 3000 transfection reagents (Life Technologies). Luciferase activity assay was performed as previously described[9, 12].

**MCT-PAH model**

PAH was induced in Sprague-Dawley rats (Charles River, NY, USA) weighing between 200-250g by a single subcutaneous injection of monocrotaline (MCT, C2401, Millipore Sigma, Oakville, ON, Canada) at a dose of 60mg/kg as previously described[13]. PBS was used as a control vehicle.

***In vivo* siRNA treatment**

Briefly, 1 nmole of siDNM2 or control-siRNA modified with orthothioates to increase stability (Integrated DNA Technology, Coralville, IA, USA) was administered on day 15, 18, 21 and 24 post-MCT injection to the anesthetized rats by nebulization in 50µl saline using an aerosol nebulizer (Kent Scientific cat #neb-1200) for lung-specific delivery.

**Hemodynamics measurement**

Pulmonary hemodynamics were assessed by right heart catheterization (RHC) on day 25 via a closed-chest approach using a 1.9F pressure-volume catheter (FTH-1912B-6018, Transonic Systems Inc., Ithaca, NY, USA) as previously described[9]. Right ventricular systolic pressure (RVSP) and cardiac output (CO) were determined using RHC.

After pulmonary hemodynamics were determined, left-ventricular end-diastolic pressure (LVEDP) was measured via a closed-chest approach from the right carotid artery. Cardiac index

(CI), mean pulmonary artery pressure (mPAP) and pulmonary vascular resistance index (PVRI) were calculated using the following formulae:

$$CI = \frac{CO}{Body\,weight\,(g)}$$

$$mPAP = 0.61\,\times\,RVSP + 2\,(mmHg)$$ [9]

$$PVRI\;=\frac{mPAP - LVEDP}{CI}$$

**Rat SU5416/hypoxia-PAH model:** SU5416/hypoxia-PAH model (Su/Hx) was created in Sprague-Dawley rats (Charles River, NY, USA) weighing between 200-250g by a single subcutaneous injection of 20 mg/kg SU5416 and housed in a hypoxic chamber with 10% oxygen, as previously described[14]. Experiments were performed 3 weeks following removal from the hypoxic chamber.

**Mouse SU5416/hypoxia-PAH model:**

Mice were placed in a normobaric chamber with an incremental decrease of oxygen levels over 3 days and were maintained for 3 weeks at 10% FiO2. Animals were given 20mg/kg SU5416 via subcutaneous injection at day 0, 7 and 14 of hypoxia. Echocardiography was performed on day 21, and right heart catheterization and tissue collection occurred on day 22.
