## Supplementary Figures for "Role of Dynamin 2 in mitochondrial fission and cell cycle regulation: *Dysregulation of a miR-124-3p-STAT3-DNM2-Drp1-RGCC pathway connects fission and cell proliferation in pulmonary arterial hypertension*"

*S1*

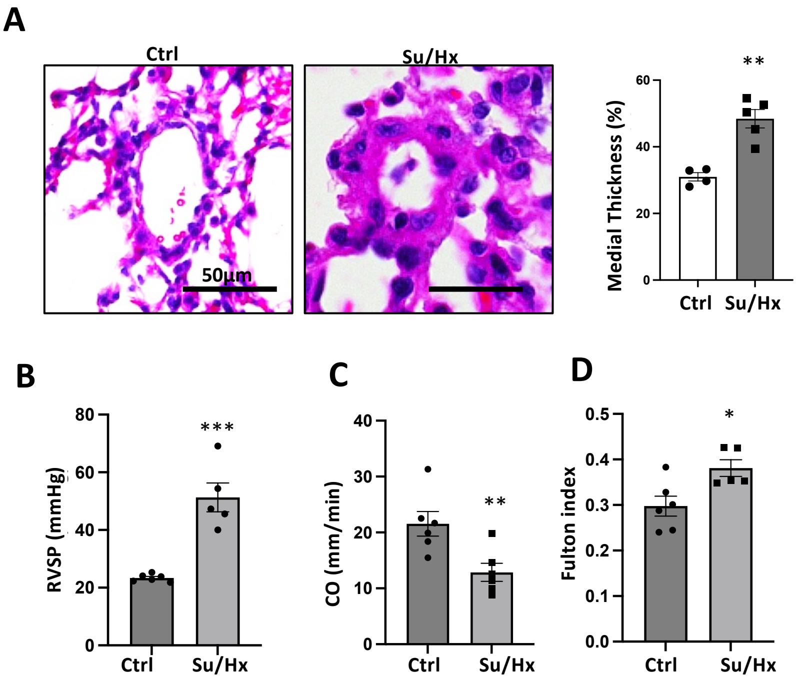


**S1: Mice treated with Su/Hx developed PAH as shown by A**) Medial hypertrophy as shown by H&E staining (n=5 mice/group). **B)** Elevated right ventricular systolic pressure (RVSP); **C)** cardiac output (CO) and **D)** fulton index (n=5/group). (**P*<0.05, ***P*<0.01, ****P*<0.001).

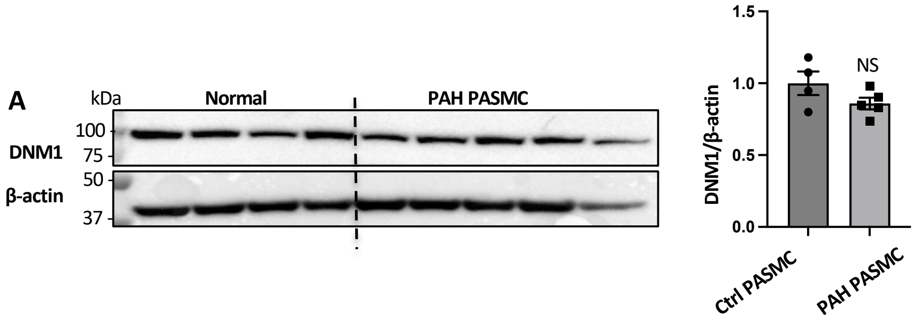


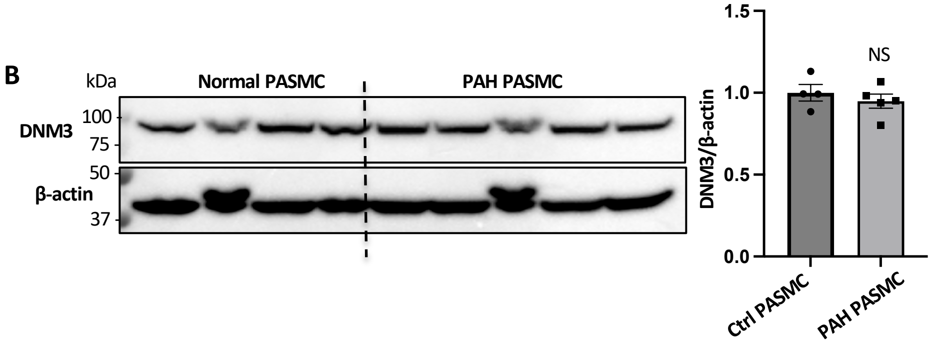


**S2: DNM1 and DNM3 are not upregulated in PAH PASMC. A-B)** Representative immunoblots and densitometries showing the expression of **A)** DNM1 and **B)** DNM3 in PAH PASMC (n=5) as compared to normal PASMC (n=4). β-actin was used as the loading control (NS, not significant).

*S3*

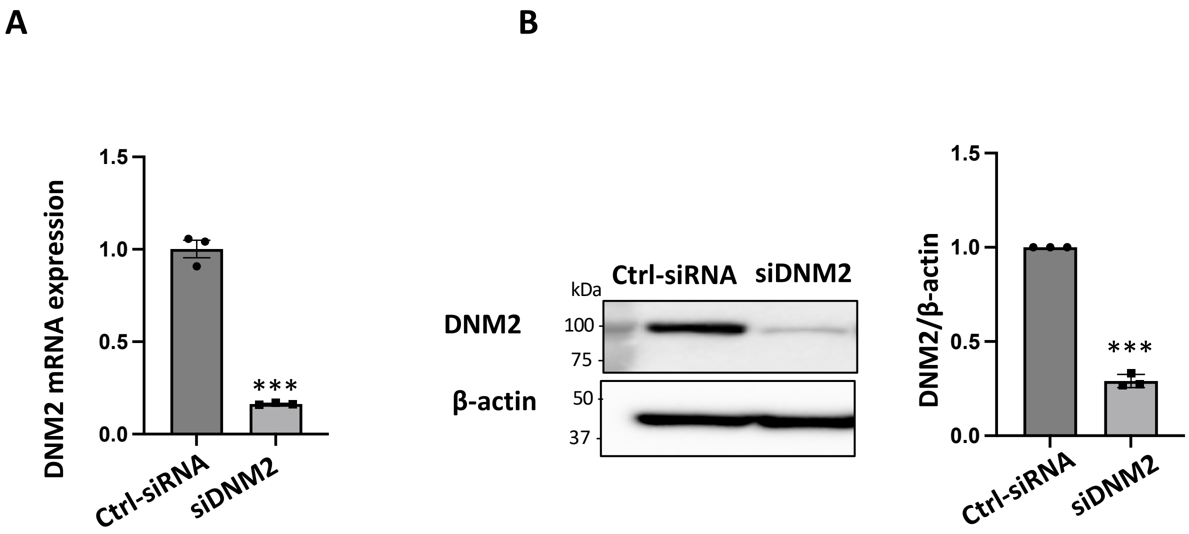


**S3: DNM2 was successfully silenced in PAH PASMC.** PAH PASMC were transfected with specified siRNA. Cells were harvested 48h following transfection and DNM2 expression was assessed using **A)** qRT PCR **B)** Representative immunoblot and densitometry. Both mRNA and protein measurements confirm successful knockdown of DNM2 (n=3 technical repeats/group, ****P*<0.001).

### PAH PASMC

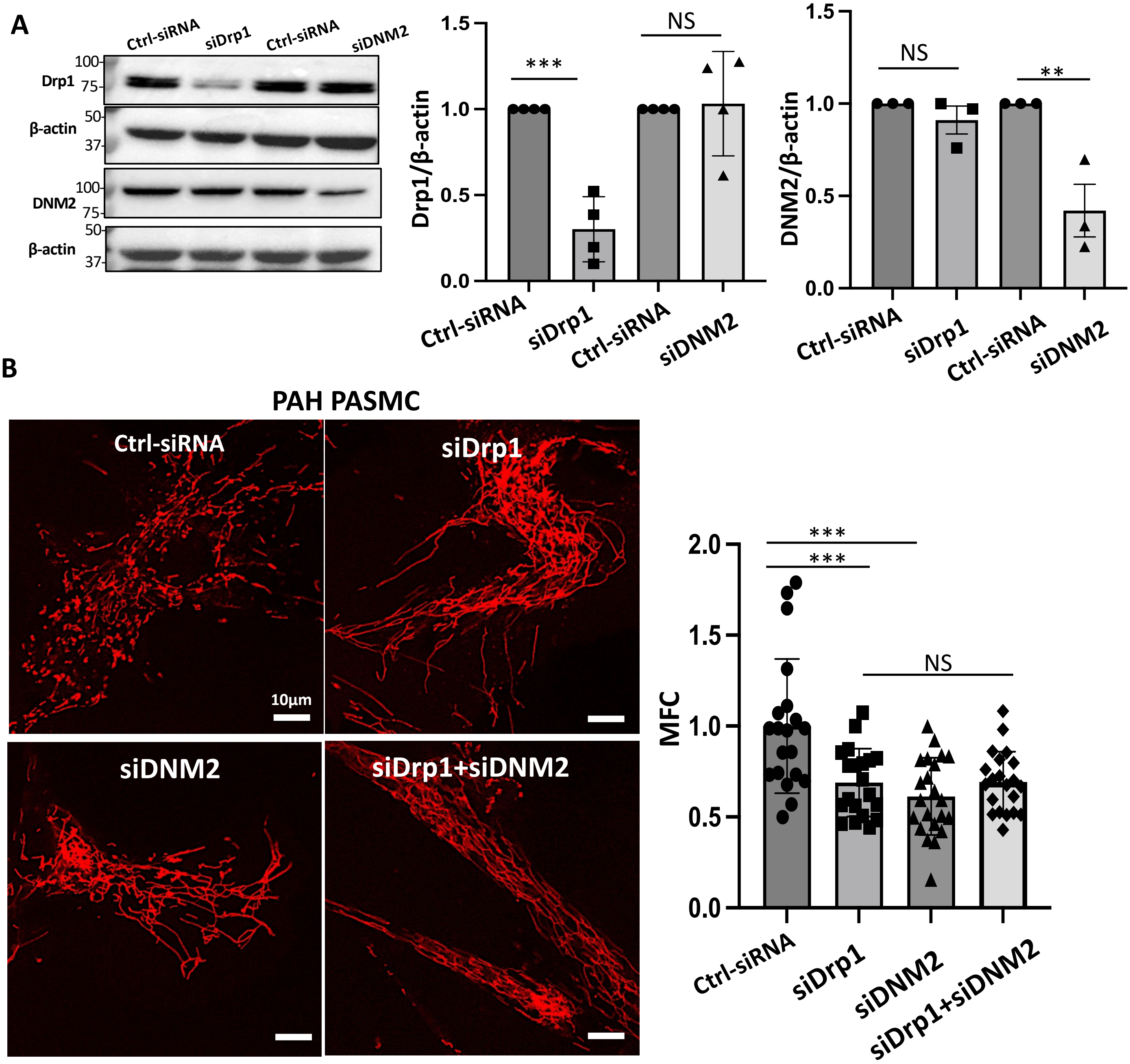


**S4: DNM2 and Drp1 expression are independent of each other and silencing both does not result in an increased inhibition of mitochondrial fission. A)** Representative immunoblot and densitometry showing that knocking down Drp1 in PAH PASMC does not alter DNM2 expression nor does knocking down DNM2 alter Drp1 expression. PAH PASMCs were transfected with specified siRNA. Immunoblot analyses were performed 48h post-transfection. β-actin was used as the loading control (n=4 PAH cell lines/group). (****P*<0.001, NS; not significant). **B)** Mitochondrial fragmentation was quantified by mitochondrial fragmentation count (MFC). PAH PASMC were transfected with specified siRNA and stained with TMRM 48h following transfection. (n=4 PAH cell lines/group). (****P*<0.001, NS; not significant).

*S5*

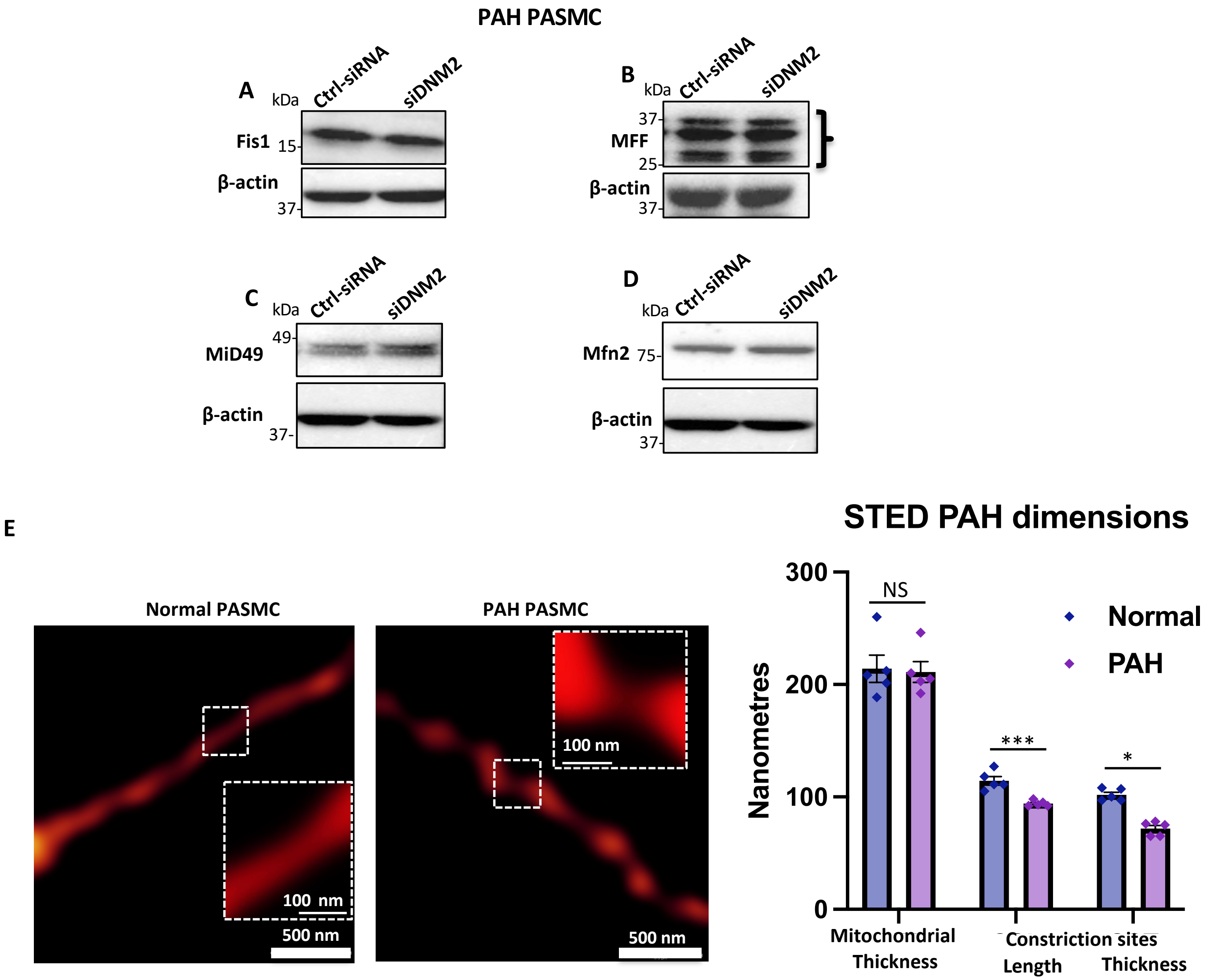


**S5: Silencing DNM2 does not alter the expression of other mitochondrial fission and fusion proteins.** Representative immunoblot showing the expression of **A)** Fis1, **B)** MFF, **C)** MiD49 and **D)** Mfn2. PAH PASMC transfected with indicated siRNA and analyzed by western blot 48h following transfection. β-actin was used as the loading control. **E) STED super resolution microscopic images showing dimensions of mitochondrial constriction sites.** Representative STED images and quantification of the dimensions of mitochondrial constriction sites in normal vs PAH PASMC. Mitochondria were stained using MitoTracker$^{TM}$ Green (n=5 cell/group) measurements for length and thickness of the mitochondrial constriction sites. Scale bar = 500 nm. (*$P < 0.05$, ***$P < 0.001$, NS, not significant).

*S6*

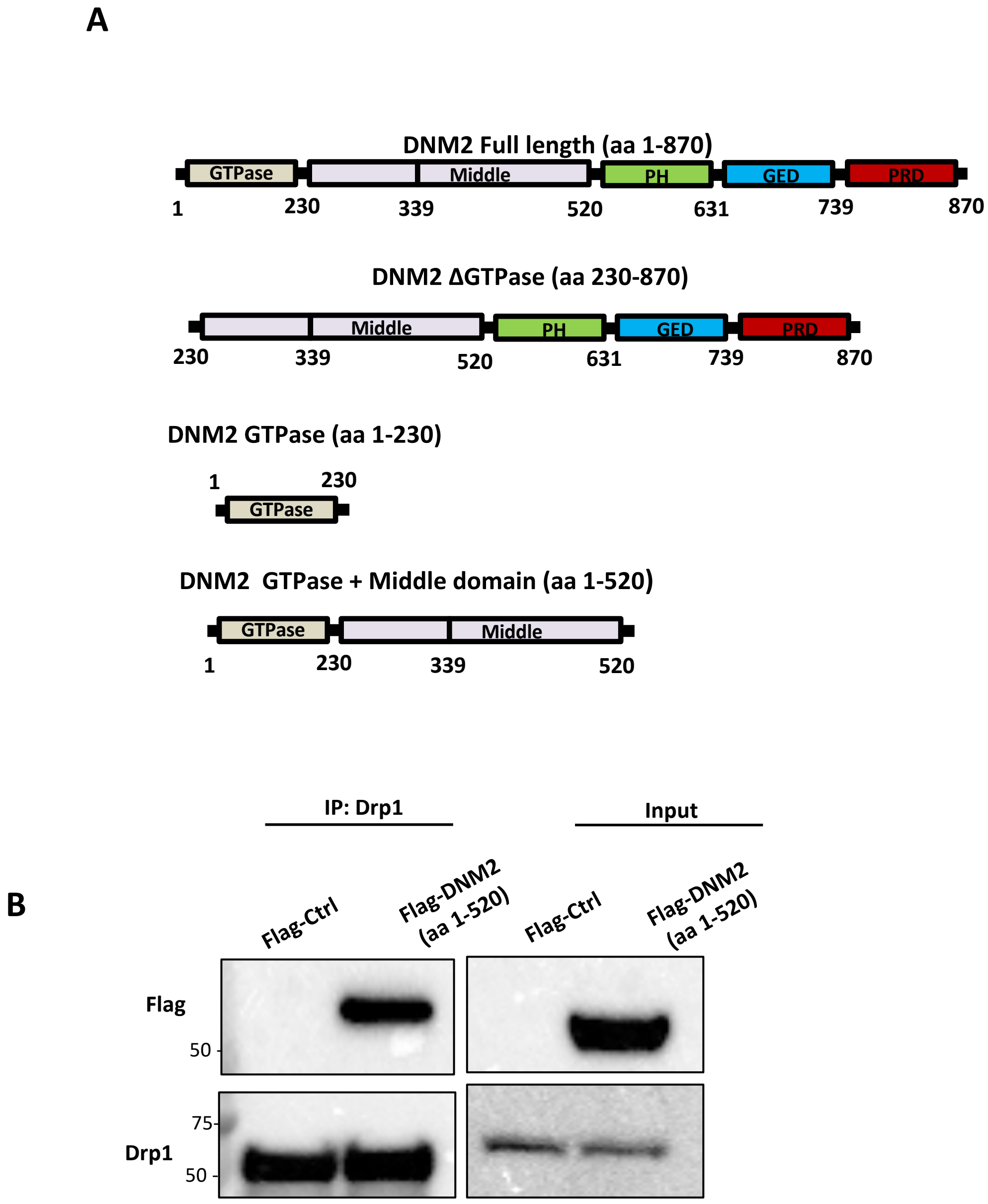


**S6: A)** A schematic representation of DNM2 full length protein, DNM2 Δ GTPase, GTPAse and GTPase + middle domain constructs showing different domains. **B)** Co-immunoprecipitation reactions showing DNM2 truncated construct containing the GTPase domain (aa 1-520) interacts with Drp1. HEK293A cells were transfected with Flag-Ctrl or Flag-DNM2 (aa 1-520) plasmids for 48 h. Co-immunoprecipitation was performed with cell lysates (2000 μg) with anti-Drp1 antibody and immunoblotted with anti-Flag antibody (top panel) and total Drp1 antibody (bottom panel). Total cell extracts (100 μg) were used as positive control for the expression of DNM2 (aa 1-520) protein and Drp1.

#### HEK293A cells

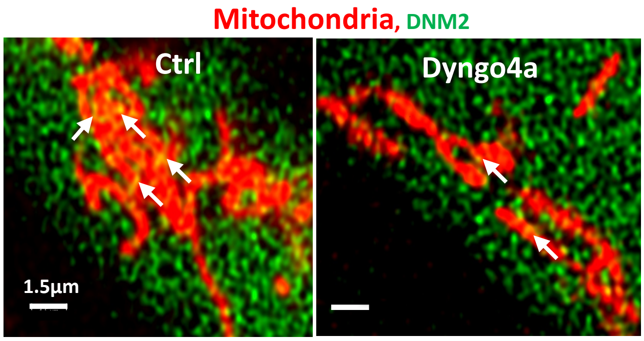


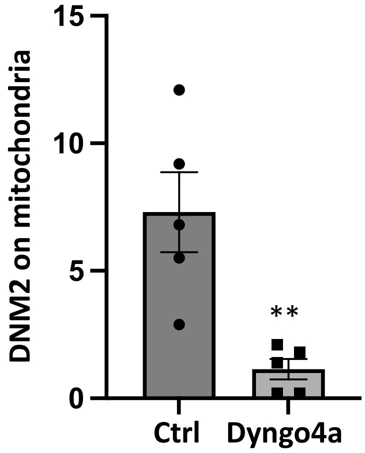


**S7: Pharmacological inhibition of GTPase activity prevented mitochondrial localization of DNM2.** HEK293A cells were transfected with DNM2-GFP plasmid (Green). The cells were treated with or without 75 µM of Dyngo4a for 24 h post-transfection. The cells were stained with MitoTracker$^{TM}$ Deep Red for mitochondria (Red). (n=5 cells/group) (**P<0.01).

**Rat MCT-PAH PASMC** **Rat Su/Hx-PAH PASMC**

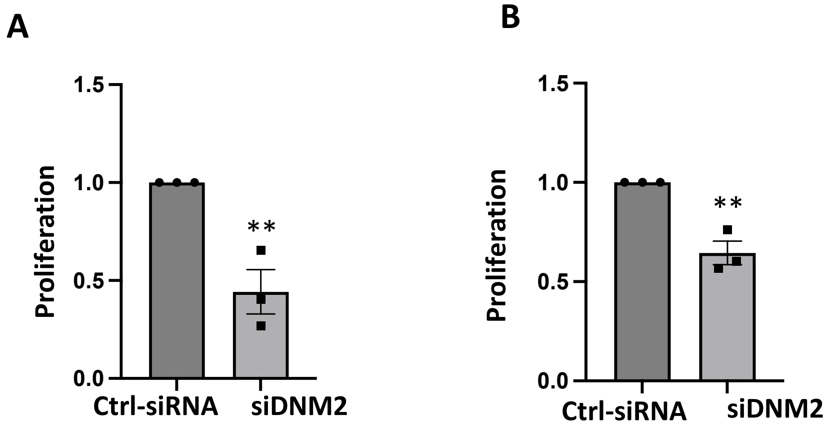


**S8: Silencing DNM2 inhibits proliferation of Rat MCT-PAH and rat Su/Hx-PAH PASMC.** The rat **A)** MCT-PAH PASMC **B)** Su/Hx-PAH PASMC were transfected with specified siRNA and EdU assay was performed 72 h post-transfection. (n=3 rat MCT-PAH cell lines /group) (**$P$<0.01)

*S9*

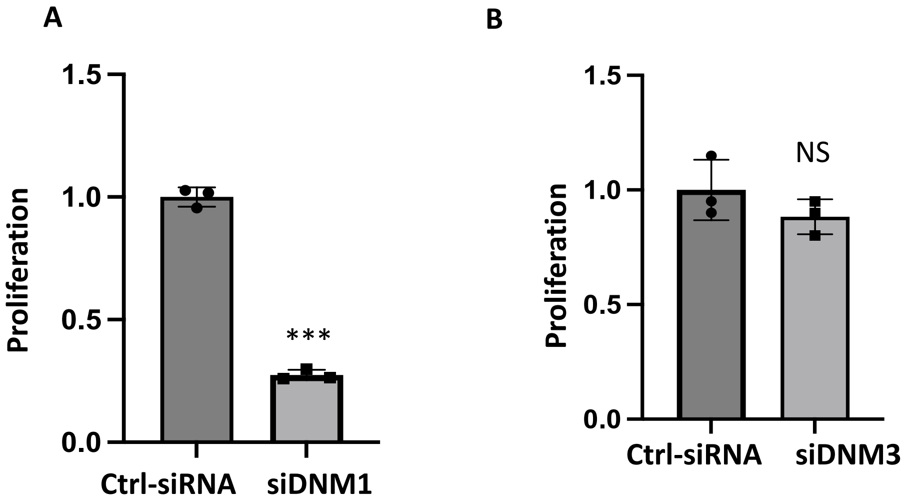


**S9: Silencing DNM1 and not DNM3 inhibits proliferation of PAH PASMC.** The PAH PASMC were transfected with **A)** siDNM1 and **B)** siDNM3. EdU assay was performed 72 h following transfection (n=3 /group. Technical repeats) (****P*<0.001, NS: not significant).

*S10*

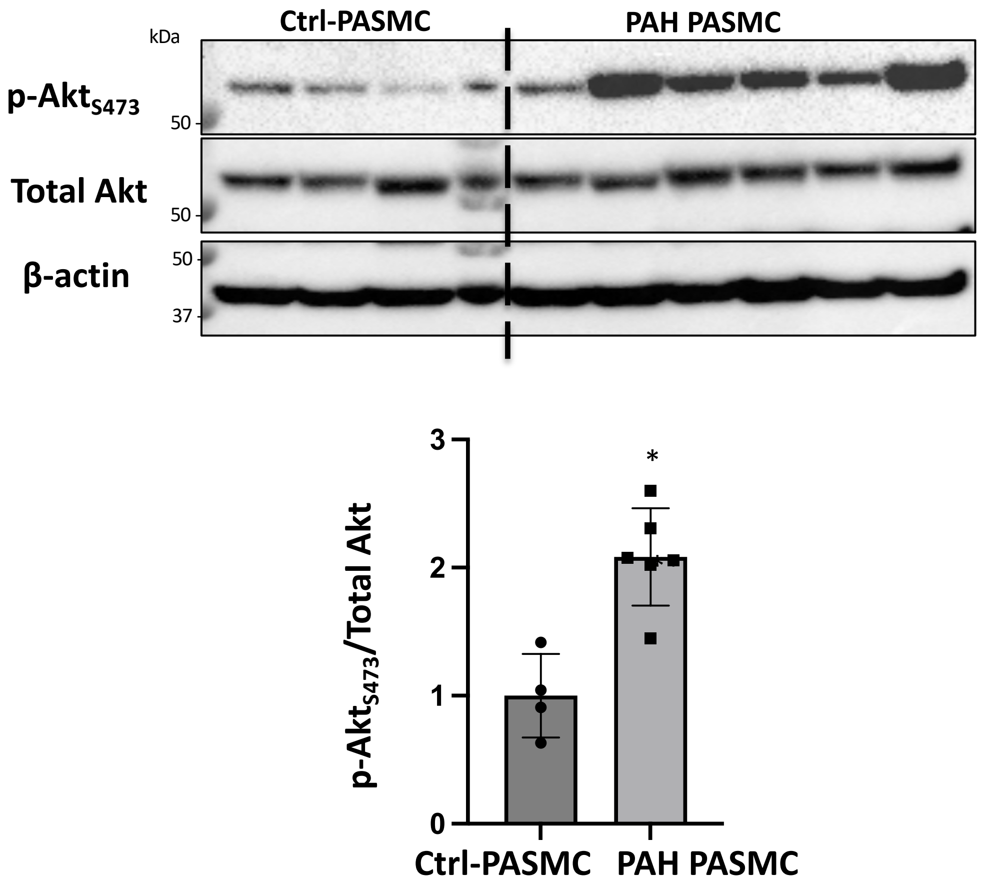


**S10: Akt is activated in PAH PASMC.** Immunoblot and densitometry performed with the cell lysates extracted from normal PASMC (n=4) and PAH PASMC (n=6). β-actin was used as the loading control. (*$P$<0.05).

***S11***

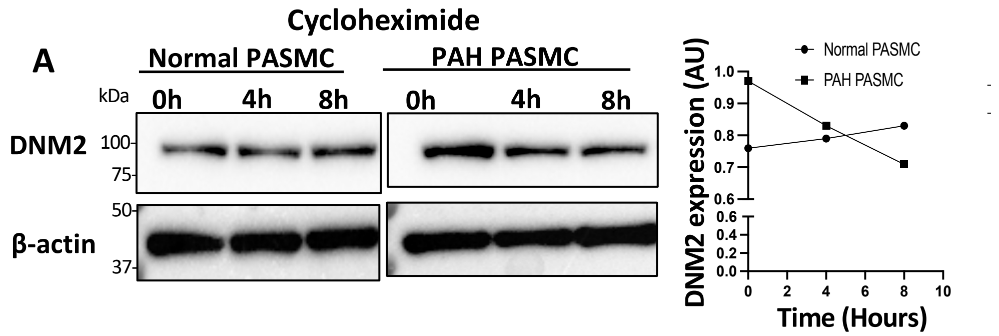


**B**

**Table. MicroRNAs predicted to bind DNM2 (TargetScan 8.0)**

| | | |
|---|---|---|
| 1. miR-454-3p | 5. miR-20-5p | 9. miR-106a-5p |
| 2. miR-130-3p | 6. miR-211-5p | 10. miR-519a-3p |
| 3. miR-301-3p | 7. miR-133a-3p | 11. miR-124-3p |
| 4. miR-17-5p | 8. miR-93-5p | 12. miR-204-5p |

Upregulated in PAH
Downregulated in PAH
Not reported in PAH

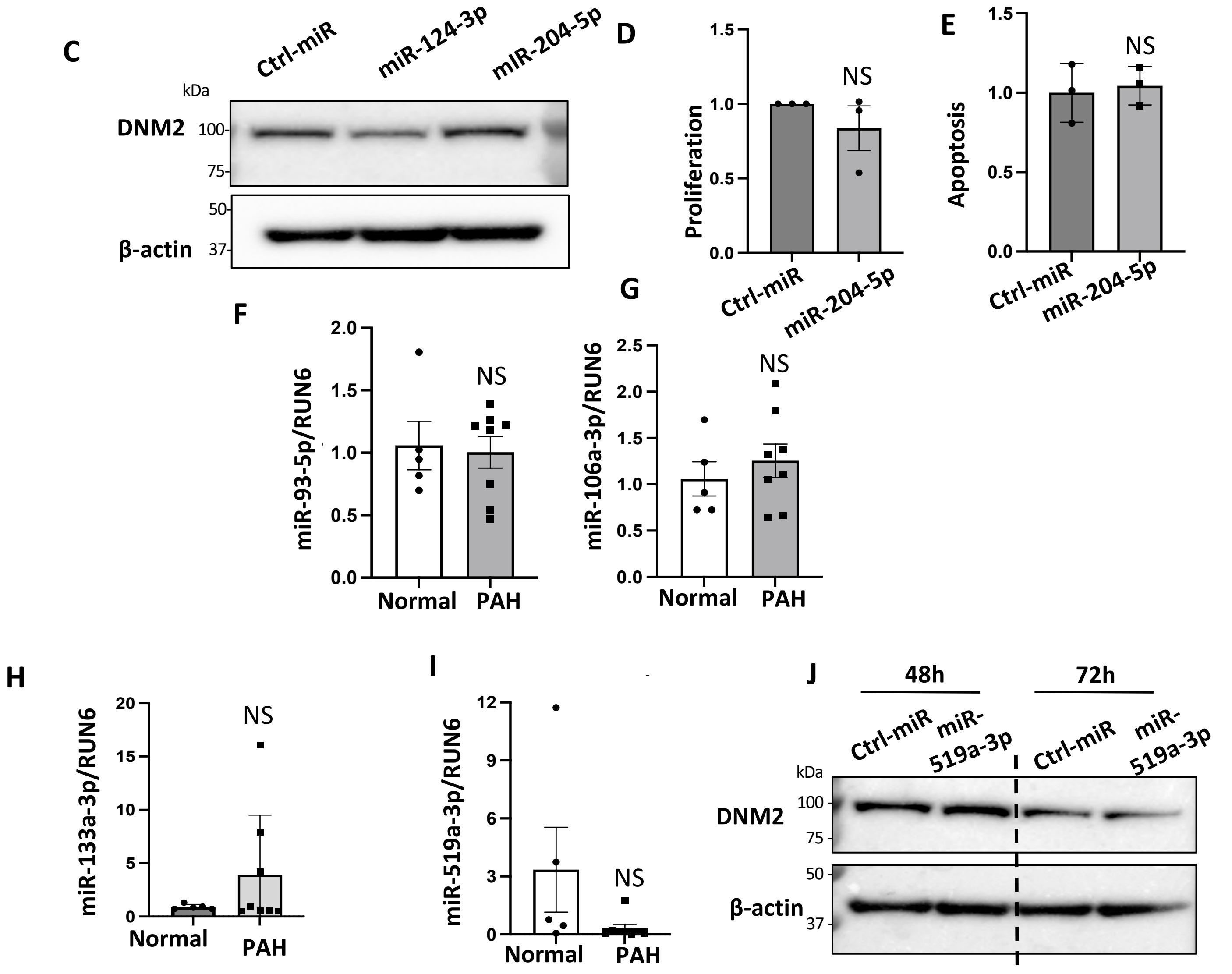

**S11: The expression of DNM2 is epigenetically regulated. A)** DNM2 protein stability is reduced in PAH PASMC: Normal and PAH PASMC were treated with cycloheximide (50µg/ml) to stop protein synthesis and the samples were collected at specified time-points. The expression of DNM2 was quantified and β-actin was used as the loading control. **B)** MicroRNAs predicted to bind DNM2 (TargetScan 8.0). Downregulated, Upregulated, Not reported miRs in PAH). **C)** Augmenting miR-204-5p does not change DNM2 expression in PAH PASMC. The cells were transfected with specified miRs for 48 h and immunoblot assay performed. β-acting was used as the loading control. **D-E)** Augmenting miR-204-5p did not **D)** inhibit proliferation or **E)** induce apoptosis in PAH PASMC. The cells were transfected with specified miRs and cell proliferation was measured by EdU incorporation assay 72 h following transfection. Apoptosis induction was determined by Annexin V + PI staining 72 h following transfection. **F-I)** qRT PCR assay showing **F)** miR-93-5p, **G)** miR-106a-5p, **H)** miR-133a-3p **I)** miR-519a-3p are not significantly downregulated in PAH PASMC. (n= 5, normal PASMC, n=8 PAH PASMC; NS, not significant). **J)** miR-519a-3p does not regulate the expression of DNM2 in PAH PASMC. Representative immunoblot showing the expression of DNM2 in PAH PASMC transfected with miR-519a-3p for 48 and 72h. β-actin was used as the loading control.

***S12***

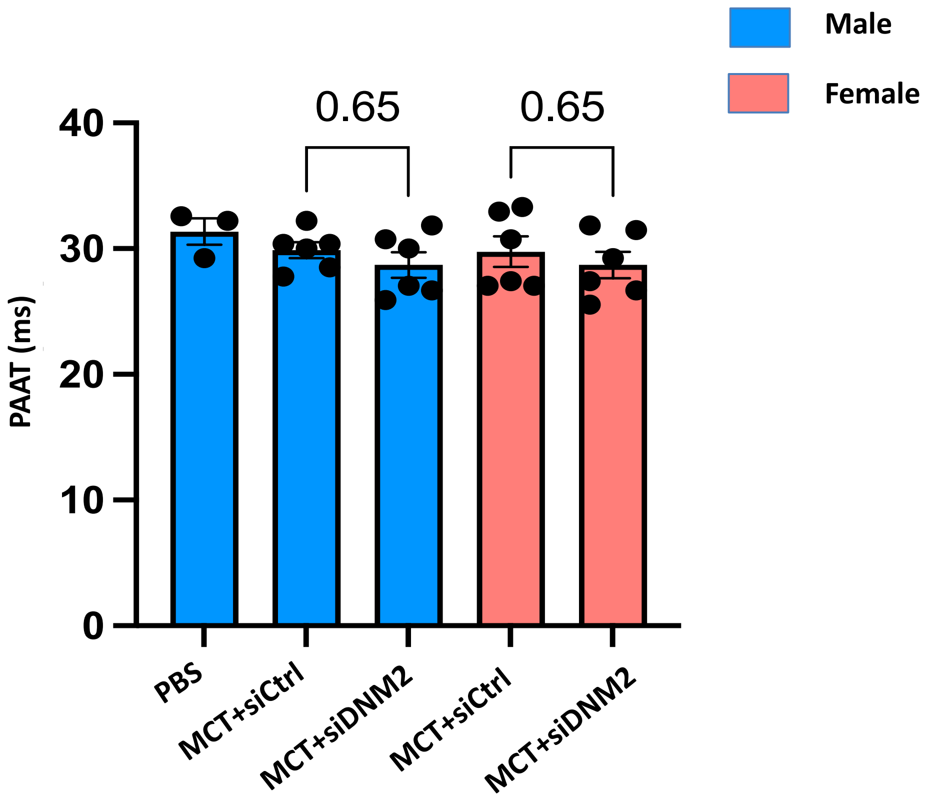


**S12: Echocardiography data on day 14 post MCT-injection.** Echocardiography on day 14 prior to nebulization shows decreased pulmonary artery acceleration time (PAAT) in MCT group. At randomization there was no significant difference in PAAT between the MCT rats that were randomly assigned to receive siCtrl vs those assigned to receive siDNM2

***S13***

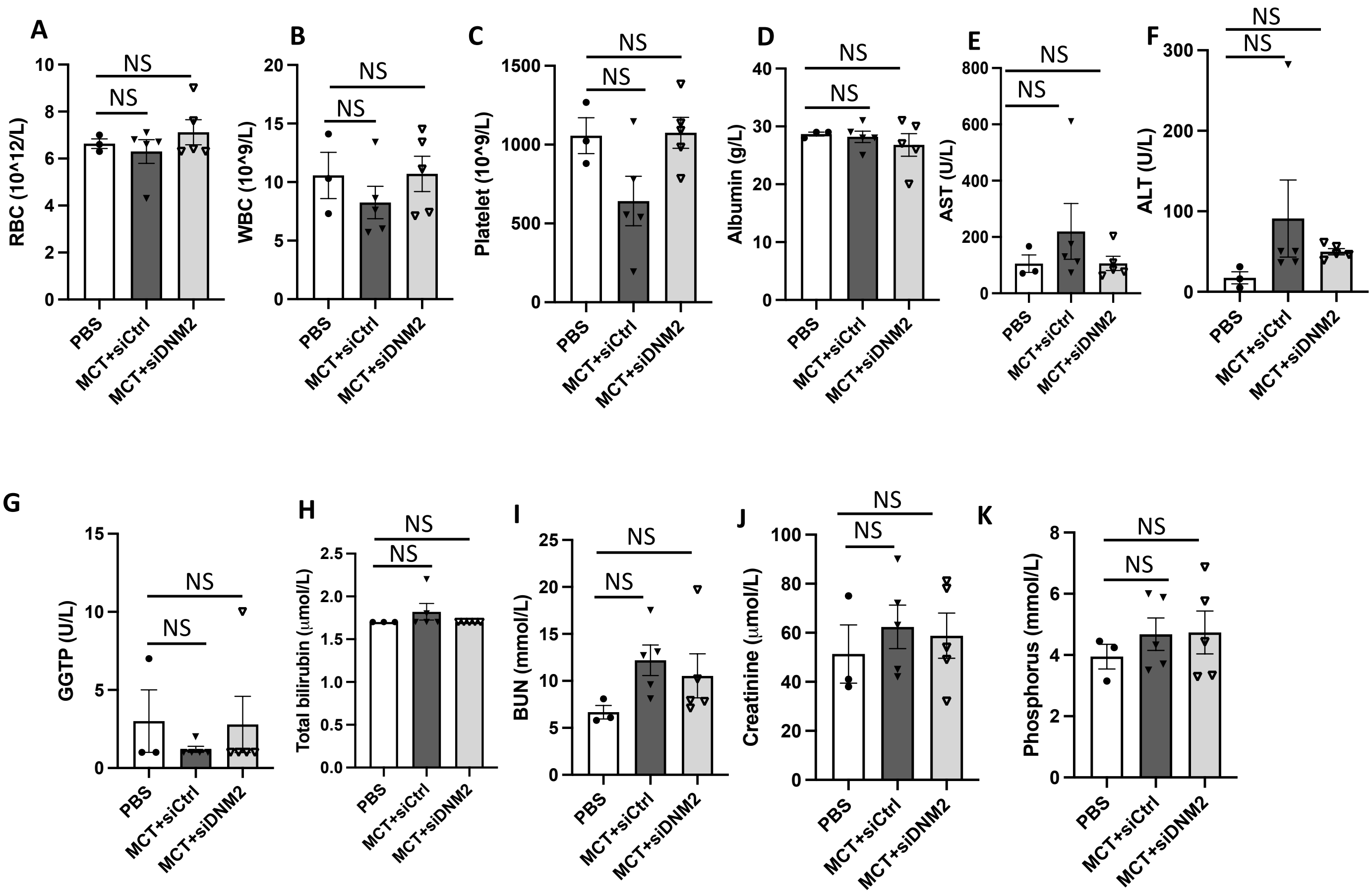


**S13: Silencing DNM2 did not induce significant hematological, hepatic and renal toxicity in MCT-PAH rats. A)** RBC: red blood cell; **B)** WBC: white blood cell; **C)** platelet; **D)** albumin; **E)** aspartate aminotransferase; (AST) **F)** alanine transaminase (ALT); **G)** The gamma-glutamyl transpeptidase (GGTP); **H)** total bilirubin; **I)** BUN: blood urea nitrogen; **J)** creatinine; **K)** phosphorus
