## Supplementary Tables for "Role of Dynamin 2 in mitochondrial fission and cell cycle regulation: *Dysregulation of a miR-124-3p-STAT3-DNM2-Drp1-RGCC pathway connects fission and cell proliferation in pulmonary arterial hypertension*"

**Supplementary Table**

**Table 1: Demographics of lung samples used for IHC to determine the expression of DNM2 in the small pulmonary arteries**

| Group | Age Range | Sex | Type |
|---|---|---|---|
| Control | 26-30 | M | - |
| Control | 31-35 | M | - |
| Control | 51-55 | M | - |
| Control | 46-50 | F | - |
| Control | 46-50 | F | - |
| PAH | 76-80 | M | Scleroderma PAH |
| PAH | 56-60 | F | IPAH |
| PAH | 71-75 | F | IPAH |
| PAH | 46-50 | F | Scleroderma PAH |
| PAH | 71-75 | F | Scleroderma PAH |

**Table 2: Top five down- and up-regulated genes in human PAH PASMC after silencing DNM2**

| Gene | $Log_2$(fold change) | log_10(adjusted p-value) |
|---|---|---|
| *DNM2* | -1.388 | 9.102 |
| *RGCC* | -1.320 | 1.662 |
| *TICAM1* | -1.235 | 2.046 |
| *RASSF3* | -1.186 | 3.395 |
| *MRPL39* | -1.140 | 1.511 |
| *IFIT1* | 0.915 | 2.053 |
| *ARL6IP1* | 0.933 | 5.792 |
| *POLR3G* | 1.078 | 3.835 |
| *CBX1* | 1.201 | 7.256 |
| *CMPK2* | 1.363 | 1.039 |

### Supplemental Table 3

| | Cluster | Category | ID | Description | p.adjust | query_size | Count | term_size | effective_domain_size | geneID | GeneRatio | BgRatio |
|---|---|---|---|---|---|---|---|---|---|---|---|---|
| GO:0000082 | query_1 | GO:BP | GO:0000082 | G1/S transition of mitotic cell cycle | 0.022357033 | 121 | 6 | 213 | 21031 | RGCC/RPS6KB1/CDK7/TMEM14B/CDKN2A/ADAMTS1 | 6/121 | 213/21031 |
| GO:2000045 | query_1 | GO:BP | GO:2000045 | regulation of G1/S transition of mitotic cell cycle | 0.027298265 | 121 | 5 | 161 | 21031 | RGCC/CDK7/TMEM14B/CDKN2A/ADAMTS1 | 5/121 | 161/21031 |
| GO:0044772 | query_1 | GO:BP | GO:0044772 | mitotic cell cycle phase transition | 0.027407585 | 4 | 2 | 432 | 21031 | DNM2/RGCC | 04-Feb | 432/21031 |
| GO:0044843 | query_1 | GO:BP | GO:0044843 | cell cycle G1/S phase transition | 0.028358563 | 121 | 6 | 241 | 21031 | RGCC/RPS6KB1/CDK7/TMEM14B/CDKN2A/ADAMTS1 | 6/121 | 241/21031 |
| GO:1903047 | query_1 | GO:BP | GO:1903047 | mitotic cell cycle process | 0.028358563 | 28 | 5 | 745 | 21031 | DNM2/RGCC/KAT2B/RPS6KB1/WRAP73 | 28-May | 745/21031 |
| GO:0044770 | query_1 | GO:BP | GO:0044770 | cell cycle phase transition | 0.031735882 | 4 | 2 | 537 | 21031 | DNM2/RGCC | 04-Feb | 537/21031 |
| GO:0000278 | query_1 | GO:BP | GO:0000278 | mitotic cell cycle | 0.032271357 | 102 | 11 | 895 | 21031 | DNM2/RGCC/KAT2B/RPS6KB1/WRAP73/NEK7/CDK7/CHMP1B/TMEM14B/CDKN2A/ZFP36L1 | 11/102 | 895/21031 |
| GO:1902806 | query_1 | GO:BP | GO:1902806 | regulation of cell cycle G1/S phase transition | 0.035042776 | 121 | 5 | 187 | 21031 | RGCC/CDK7/TMEM14B/CDKN2A/ADAMTS1 | 5/121 | 187/21031 |
| GO:0000086 | query_1 | GO:BP | GO:0000086 | G2/M transition of mitotic cell cycle | 0.040175575 | 1 | 1 | 138 | 21031 | DNM2 | 01-Jan | 138/21031 |
| GO:0044839 | query_1 | GO:BP | GO:0044839 | cell cycle G2/M phase transition | 0.041789124 | 1 | 1 | 154 | 21031 | DNM2 | 01-Jan | 154/21031 |
| GO:1900087 | query_1 | GO:BP | GO:1900087 | positive regulation of G1/S transition of mitotic cell cycle | 0.04343418 | 4 | 1 | 41 | 21031 | RGCC | 04-Jan | 41/21031 |
| GO:0007346 | query_1 | GO:BP | GO:0007346 | regulation of mitotic cell cycle | 0.043605307 | 121 | 8 | 496 | 21031 | RGCC/RPS6KB1/NEK7/CDK7/TMEM14B/CDKN2A/ZFP36L1/ADAMTS1 | 8/121 | 496/21031 |
| GO:0022402 | query_1 | GO:BP | GO:0022402 | cell cycle process | 0.044288724 | 30 | 6 | 1276 | 21031 | DNM2/RGCC/KAT2B/RPS6KB1/WRAP73/NUP37 | 30-Jun | 1276/21031 |
| GO:0045787 | query_1 | GO:BP | GO:0045787 | positive regulation of cell cycle | 0.045647508 | 26 | 3 | 349 | 21031 | RGCC/KAT2B/RPS6KB1 | 26-Mar | 349/21031 |
| GO:0045931 | query_1 | GO:BP | GO:0045931 | positive regulation of mitotic cell cycle | 0.046711227 | 26 | 2 | 120 | 21031 | RGCC/RPS6KB1 | 26-Feb | 120/21031 |
| GO:1902808 | query_1 | GO:BP | GO:1902808 | positive regulation of cell cycle G1/S phase transition | 0.04856887 | 4 | 1 | 56 | 21031 | RGCC | 04-Jan | 56/21031 |
| GO:1901990 | query_1 | GO:BP | GO:1901990 | regulation of mitotic cell cycle phase transition | 0.052362707 | 121 | 6 | 332 | 21031 | RGCC/CDK7/TMEM14B/CDKN2A/ZFP36L1/ADAMTS1 | 6/121 | 332/21031 |
| GO:1901992 | query_1 | GO:BP | GO:1901992 | positive regulation of mitotic cell cycle phase transition | 0.060605754 | 4 | 1 | 89 | 21031 | RGCC | 04-Jan | 89/21031 |
| GO:1901989 | query_1 | GO:BP | GO:1901989 | positive regulation of cell cycle phase transition | 0.066675094 | 4 | 1 | 112 | 21031 | RGCC | 04-Jan | 112/21031 |
| GO:0010564 | query_1 | GO:BP | GO:0010564 | regulation of cell cycle process | 0.070052539 | 121 | 9 | 724 | 21031 | RGCC/KAT2B/CDK7/CHMP1B/TMEM14B/CDKN2A/TXLNG/ZFP36L1/ADAMTS1 | 9/121 | 724/21031 |
| GO:0051726 | query_1 | GO:BP | GO:0051726 | regulation of cell cycle | 0.071705435 | 121 | 12 | 1105 | 21031 | RGCC/KAT2B/RPS6KB1/NEK7/CDK7/CHMP1B/TMEM14B/CDKN2A/TXLNG/ZFP36L1/HIPK2/ADAMT | 12/121 | 1105/21031 |
| GO:0090068 | query_1 | GO:BP | GO:0090068 | positive regulation of cell cycle process | 0.072608547 | 21 | 2 | 257 | 21031 | RGCC/KAT2B | 21-Feb | 257/21031 |
| GO:1901991 | query_1 | GO:BP | GO:1901991 | negative regulation of mitotic cell cycle phase transition | 0.080692916 | 4 | 1 | 181 | 21031 | RGCC | 04-Jan | 181/21031 |
| GO:0010948 | query_1 | GO:BP | GO:0010948 | negative regulation of cell cycle process | 0.082425315 | 21 | 2 | 302 | 21031 | RGCC/KAT2B | 21-Feb | 302/21031 |
| GO:1901987 | query_1 | GO:BP | GO:1901987 | regulation of cell cycle phase transition | 0.086082511 | 121 | 6 | 433 | 21031 | RGCC/CDK7/TMEM14B/CDKN2A/ZFP36L1/ADAMTS1 | 6/121 | 433/21031 |
| GO:0007049 | query_1 | GO:BP | GO:0007049 | cell cycle | 0.086774793 | 4 | 2 | 1819 | 21031 | DNM2/RGCC | 04-Feb | 1819/21031 |
| GO:0045930 | query_1 | GO:BP | GO:0045930 | negative regulation of mitotic cell cycle | 0.090306023 | 4 | 1 | 232 | 21031 | RGCC | 04-Jan | 232/21031 |
| GO:1901988 | query_1 | GO:BP | GO:1901988 | negative regulation of cell cycle phase transition | 0.095683283 | 4 | 1 | 260 | 21031 | RGCC | 04-Jan | 260/21031 |
